# Proteomics and personalized patient-derived xenograft models identify treatment opportunities for a progressive malignancy within a clinically actionable timeframe and change care

**DOI:** 10.1101/2024.07.04.24309923

**Authors:** Georgina D. Barnabas, Tariq A. Bhat, Verena Goebeler, Pascal Leclair, Nadine Azzam, Nicole Melong, Colleen Anderson, Alexis Gom, Seohee An, Enes K. Ergin, Yaoqing Shen, Andy J. Mungall, Karen L. Mungall, Christopher A. Maxwell, Gregor S.D. Reid, Martin Hirst, Steven Jones, Jennifer A. Chan, Donna L. Senger, Jason N. Berman, Seth J. Parker, Jonathan W. Bush, Caron Strahlendorf, Rebecca J. Deyell, Chinten J. Lim, Philipp F. Lange, PROFYLE Program

## Abstract

Increased access to high-throughput DNA sequencing platforms has transformed the diagnostic landscape of pediatric malignancies by identifying and integrating actionable genomic or transcriptional features that refine diagnosis, classification, and treatment. Yet less than 10% of treated patients show a positive response and translating precision oncology data into feasible and effective therapies for hard-to-cure childhood, adolescent, and young adult malignancies remains a significant challenge. Combining the identification of therapeutic targets at the protein and pathway levels with demonstration of treatment response in personalized models holds great promise. Here we present the case for combining proteomics with patient-derived xenograft (PDX) models to identify personalized treatment options that were not apparent at genomic and transcriptomic levels. Proteome analysis with immunohistochemistry (IHC) validation of formalin-fixed paraffin-embedded sections from an adolescent with primary and metastatic spindle epithelial tumor with thymus-like elements (SETTLE) was completed within two weeks of biopsy.

The results identified an elevated protein level of SHMT2 as a possible target for therapy with the commercially available anti-depressant sertraline. Within 2 months and ahead of a molecular tumor board, we confirmed a positive drug response in a personalized chick chorioallantoic membrane (CAM) model of the SETTLE tumor (CAM-PDX). Following the failure of cytotoxic chemotherapy and second-line therapy, a treatment of sertraline was initiated for the patient. After 3 months of sertraline treatment the patient showed decreased tumor growth rates, albeit with clinically progressive disease.

Significance: Overall, we demonstrate that proteomics and fast-track personalized xenograft models can provide supportive pre-clinical data in a clinically meaningful timeframe to support medical decision-making and impact the clinical practice. By this we show that proteome-guided and functional precision oncology are feasible and valuable complements to the current genome-driven precision oncology practices.

## Introduction

Genome-driven precision medicine revolutionized cancer treatment by tailoring therapies to individual tumors’ molecular profiles, rather than the conventional generic approach. Utilizing genomic strategies and clinical data, precision medicine strives for more targeted and less toxic therapies, potentially enhancing outcomes for cancer patients (*1, 2*). Yet, while actionable genomic or transcriptional level changes are observed in >70% of pediatric and young adult patients with solid tumors, half of patients treated with a targeted therapy show a positive response (*3–5*). Genome-driven precision oncology maps the causal alterations, while proteomics provides a quantitative assessment of cellular processes providing complementary insight closer to the disease and therapy response phenotype. Proteome-guided precision oncology further enables a direct assessment of the drug target levels as well as the status of downstream resistance mechanisms. Though still at an early stage, integrating global proteomic analysis into precision oncology holds promise, particularly for personalized target identification (*6, 7*).

Despite therapeutic and diagnostic advancements, cancer remains a leading cause of mortality in children, adolescents, and young adults (CAYA), with an average of 943 new cases and 119 deaths annually in Canada (*8, 9*). The molecular landscape of CAYA cancers differs significantly from that of adult malignancies, posing challenges in translating precision oncology data into effective therapies (*10, 11*). Genome-driven precision oncology trials focussing on CAYA patients have been initiated after their adult counterparts and only recently started to report initial findings. For example, in the MAPPYACTS trial, 56% of children had actionable alterations, but only 16% of those receiving targeted therapy (19%) had partial or complete response (*12*). The INFORM registry reported 48% of cases with actionable alterations, 16% receiving targeted therapy, and no reports on the response or survival status (*13*).

PRecision Oncology For Young peopLE (PROFYLE) is a collaborative pan-Canadian project aiming to provide more effective treatment options to CAYA with advanced cancers. In parallel to genome-driven precision oncology, the prospect of proteome-guided and functional precision oncology approaches are explored. Murine patient-derived xenografts (PDX) represent *in vivo* tumor characteristics (*14*) for assessing potential treatments. However, effective xenograft modeling of pediatric solid tumors has to overcome challenges arising from the limited quantity and of available viably cryopreserved tissues that frequently limit the utility of the traditional immune-incompetent mouse models (*15*). A faster alternative is the embryonic chick chorioallantoic membrane (CAM), a highly vascularized *in vivo* model with an underdeveloped immune system, which provides a suitable microenvironment for growth and drug response assessments of tumor xenografts (*16–19*). PDX models created using larval zebrafish have also become invaluable for pre-clinical cancer research due to their high reproductive rate, lack of a fully adaptive immune system and cost-effectiveness (*20, 21*). Both the CAM and zebrafish models are established as robust and readily accessible *in vivo* systems, distinguished by their rapid experimental turnover.

Target selection in precision oncology is often facilitated by the identification of molecular similarity to patient cohorts with well-established targeted therapies. To evaluate if proteome-guided precision oncology can provide de-novo target prioritization in the absence of molecular grouping, we opted to study a rare and challenging CAYA malignancy, a spindle epithelial tumor with thymus-like elements (SETTLE) with progressive disease. SETTLE originates from ectopic thymus tissue or branchial pouch remnants in the thyroid and presents a distinct biphasic pattern of spindle and epithelial cells. Although SETTLE is categorized as a slow growing malignancy with a 5-year overall survival of >80%, metastasis have been reported in up to 41% of patients with at least 5-year follow up, with a latency period of up to 10 years (*22, 23*). Molecular pathogenesis and clinical outcomes of SETTLE remain poorly understood due to limited case studies (*24*), which provide valuable clinical and histological insights but lack comprehensive molecular characterization. We hypothesized that quantitative whole proteome profiling and drug response validation using PDX models could provide on-time pre-clinical insight complementary to whole exome and transcriptome sequencing and inform clinical decision-making.

Here, we report the integration of proteomics with PDX models revealing potential personalized treatment options that were not evident from genome and transcriptome analyses. Following rapid target identification by comprehensive mass spectrometry proteomics and validation by immunohistochemistry, we confirmed response to prioritized SHMT2-targeting sertraline treatment *in vitro* from murine PDX cells, and *in vivo* with CAM and larval zebrafish PDXs. Metabolic tracing confirms serine addiction of SETTLE cells and validates on-target effect of sertraline. This study underscores the value of combining proteomics with personalized xenograft models in pre-clinical research to inform medical decisions and influence clinical practice for the individual patient, and also encourages accelerated integration of proteome and functional approaches into precision oncology pipelines.

## Results

### Patient history

The child had previously presented with asymptomatic right neck swelling, which had been incidentally noted. At initial diagnosis, an ultrasound showed a 3.8 x 2.3 x 4.5cm heterogeneous soft tissue mass replacing the right thyroid lobe, with no associated adenopathy. The patient had already undergone a right hemithyroidectomy for the disease and was confirmed as a SETTLE tumor as reported (*25*). This diagnosis started ongoing cycles of testing, treatment, including genome profiling-guided targeted therapy, period of remission, and recurrence. The patient was experiencing progressive disease when enrolled in the PROFYLE study.

We conducted LC-MS/MS-based quantitative whole proteome profiling, candidate target identification, and IHC target validation within 2 weeks of biopsy. *In PDX* drug response evaluation was conducted in parallel to whole exome/transcriptome sequencing within the 2 months leading up to the case evaluation by the PROFYLE molecular tumor board and subsequent initiation of innovative therapy (Fig. 1A, B).

**Figure 1:**
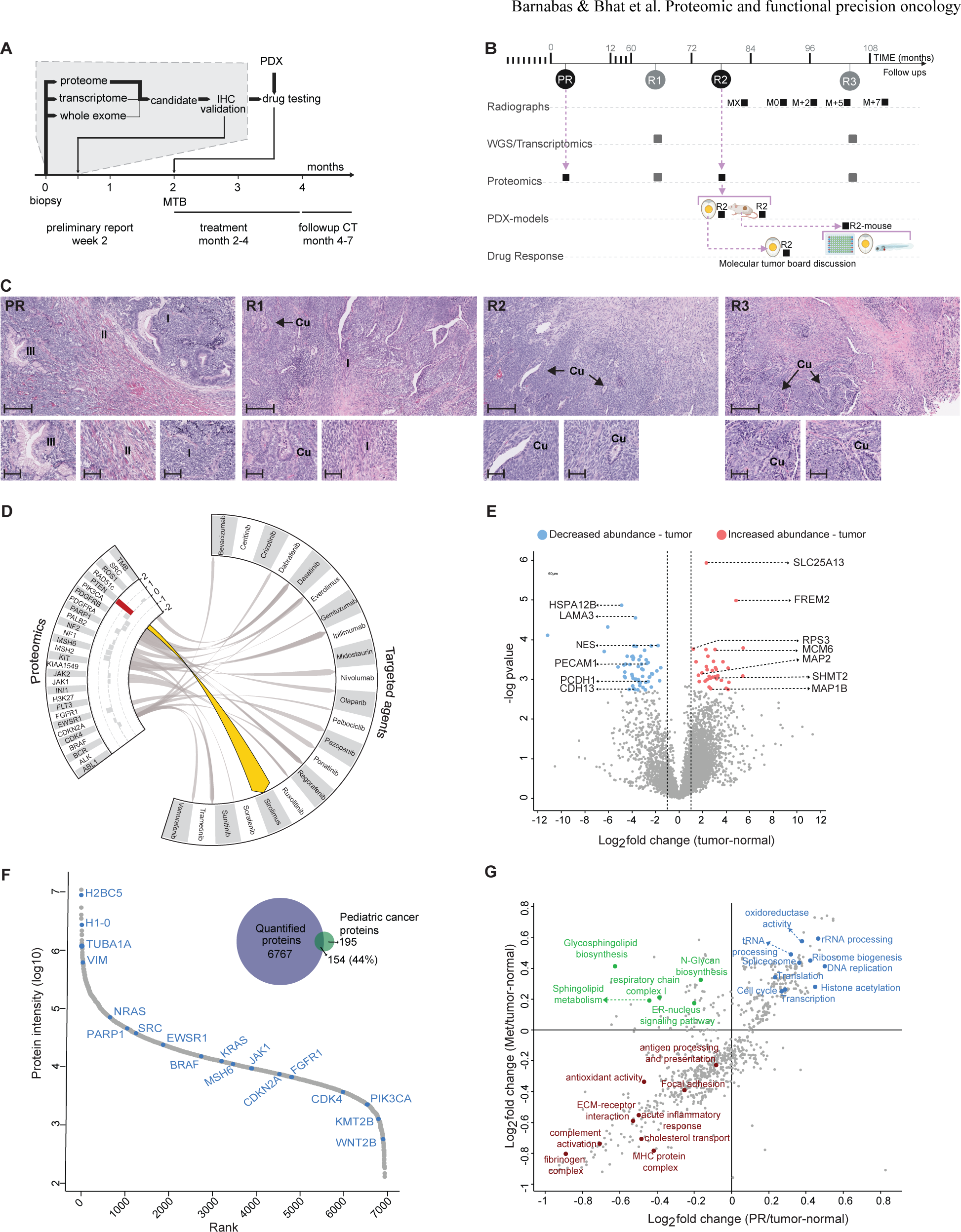
Multi-omics molecular profiling of a progressive SETTLE case. **A.** Timeline of precision diagnostics for the SETTLE disease course, including multi-omics molecular tumor profiling, real-time target identification, and validation using personalized xenograft models in providing timely pre-clinical support for medical decision-making. **B.** Patient journey from the resection of the tumor (PR) through three distant recurrences (R1, R2, R3). The molecular analyses, radiographs (Months at time X, MX to M+7), PDX models, and drug sensitivity assays conducted are linked to the corresponding time points. **C.** H&E stained sections of the primary resection (PR) and relapse biopsies (R1-R3). PR demonstrates a biphasic mesenchymal pattern with heterologous elements including gastric foveolar-lined glands showing three patterns, a small blue cell population with a fascicular growth pattern (I). A more collagenous and sclerotic pattern (II). The heterologous epithelial elements show foveolar-like epithelium (III). R1 shows a similar pattern to the resection specimen, including a hypocellular area with increased collagen which transitions into a hypercellular spindle cell area. The cellular nests and fascicles with more collagenous areas (I). Areas of cuboidal-lined (Cu) epithelium are seen. R2 and R3 show hypocellular collagenous tumor transitioning to hypercellular spindled areas with cuboidal-lined (Cu) epithelium. Scale bars: 200 µm in main image; 60 µm in insets. **D.** Proteome changes do not suggest sensitivity to 21 established therapies. Circular plot showing the targeted proteomic analysis focusing on proteins associated with routinely used therapies in the third lung relapse (R3). Log2 fold-change values, calculated from the tumor vs normal comparison, are displayed on the proteomics side of the plot. grey: non-significant changes and associations; red arrow: proteome change counter indicative of drug sensitivity. **E.** Volcano plot showing the global analysis of proteome perturbation between R3 and adjacent normal regions. Proteins significant with student’s t-test at 0.01 FDR and with log2 fold change >1 are highlighted in red for increased abundance in tumor and in blue for reduced abundance in tumor. **F.** Quantified proteins ranked by their average intensity across all samples to evaluate the dynamic range and depth of the proteome coverage. Venn diagram showing the overlap of all proteins quantified in this study with known pediatric cancer-associated proteins. **G.** 2D-enrichment analyses of log2 fold changes between primary resection vs adjacent normal tissue and those between lung relapses vs corresponding adjacent normal tissues against GO terms using Fisher-exact test (FDR 0.05%).

### Genomic and transcriptomic profiles at first relapse (R1)

On a routine follow-up visit, a chest x-ray showed multiple bilateral pulmonary nodules. A re-staging chest computed tomography (CT) showed multiple, bilateral pulmonary nodules up to 20 mm with no change in the right hemithyroidectomy surgical bed. A positron emission tomography/CT showed mildly avid pulmonary nodules, but no local recurrence or other distant metastatic disease. An infrared-guided video-assisted thoracoscopic surgery wedge resection of a pulmonary nodule confirmed SETTLE recurrence.

Whole genome analysis of the thoracoscopic biopsy of relapse (R1) showed a heterozygous variant of uncertain significance (VUS) p.L780F in FLT3 and a subclonal VUS in MUC4. The genome appeared to be stable with no indication of microsatellite instability or homologous recombination deficiency, and no detectable somatic copy number variation or loss of heterozygosity that affected coding regions. Multiple cancer-related genes showed high expression in the biopsy, including ROS1, FGFR1, EGFR, and PDGFA in kinase signaling, SMO, GLI2, and GLI3 in the hedgehog pathway, as well as IRS4, HES4, and MYCN. Among those involved in cell cycle regulation, MDM4 showed high expression while CDKN1A had low expression (Suppl Table 1A). Based on the genomic finding, the patient was treated with Sorafenib for 20 months. The tumour remained stable until detection of further progression.

### Multi-omics molecular profiling of relapse (R3)

Following the lack of response to conventional chemotherapy and acquired resistance to receptor tyrosine kinase inhibitor, the criteria for progressive disease was attained. Additional pulmonary resection (Fig. 1C) was performed and followed up by multi-omics analyses, for the first time including proteome analysis, to identify additional molecular targets and therapeutic options. Whole exome sequencing and RNAseq analysis of fresh frozen samples from new thoracoscopic biopsies of pulmonary nodules (R3) revealed tumor genome evolution since R1. In particular, R3 showed increased mutational burden (0.77 mut/Mb) compared to R1 (0.37 mut/Mb) and increased presentation of mutation signature after treatment versus R1. R3 carried the same notable variants previously identified in R1. These included FLT3 p.L780F, a copy loss and truncating mutation in ARID1A:p.R1335*, a subclonal mutation in SF3B1:p.K700E, and a copy loss in MUS81. Gene expression levels were similar between R1 and R3 biopsies, but a few showed higher expression in R3 compared to R1 including the cancer-related genes SPINK1, MMP8, HOXB13, IGF2, HAVCR2, CA9, and MET (Suppl Table 1B). Potential options to target the new findings were discussed at the molecular tumor board, but none were deemed clinically actionable.

### Proteomics reveals potentially actionable targets of SETTLE

In parallel to the sequencing analysis, whole proteome profiling (R3) was seamlessly interfaced with routine pathology workflows. Immediately following the biopsy, the anatomical pathology laboratory at BC Children’s Hospital prepared H&E-stained 10 µm FFPE sections from clinical FFPE blocks and isolated the tumor and adjacent normal regions using macro-dissection. Following transfer to the research laboratory, we used rapid automated sonication-free acid-assisted proteome (ASAP) processing (*26*) and data-independent acquisition (DIA) liquid chromatography-mass spectrometry (LC-MS/MS) to assemble quantitative proteome maps comparing the abundance of 4703 proteins in metastatic nodules and adjacent normal lung regions.

We first conducted a targeted analysis of the proteome maps focusing on proteins associated with routinely used therapies. None of the 14 proteins linked to the 21 targeted agents considered showed significant alterations (Fig. 1D) that would have supported their prioritization. The only significant change was an increased abundance of PTEN in tumor nodules, which is generally linked to a tumor suppressive effect (Fig. 1D) (*27*). The increased abundance of PTEN may serve as a counterindication for using Sirolimus, which is commonly prioritized for tumors with PTEN copy loss. Considering the lack of proteome changes supporting established therapies, we investigated the global proteome perturbation and found downregulation of cell adhesion proteins (CDH13, PCDH1) and upregulation of proteins involved in microtubule assembly (MAP1B, MAP2), cell cycle (MCM6), amino acid transport (SLC25A13) and serine metabolism (SHMT2) (Fig. 1E). Importantly, upregulation of SHMT2 was suggestive of serine addiction with possible therapeutic options.

To solidify the hypothesis of serine addiction and to gain more insights into the dysregulation of proteins and pathways in SETTLE, we expanded the global proteome profiling to all available tumor samples, including the primary tumor and earlier lung metastases (R1 and R2). Because we worked with FFPE tissues, it was straightforward to retrieve archival tissue sections from all earlier time points within days. Across the primary tumor, all lung metastases, and adjacent normal regions, we quantified 6921 proteins covering an extensive abundance range of five orders of magnitude, demonstrating in-depth proteome coverage (Fig. 1F). Among these, we quantified 154 of a set of 349 cancer-associated proteins that we had previously assembled with a focus on drivers and therapeutic targets in pediatric malignancies (*28*), confirming comprehensive coverage of the most relevant drivers and pathways (Fig. 1F). The median coefficient of variation (CV) between sequential sections was between 8-32% with a median of 16% (Suppl Fig. 1A).

Differential analysis between tumors and adjacent normal regions showed 795 upregulated proteins and 728 downregulated proteins in primary tumor and lung metastases (Suppl Fig. 1B). Pathway analysis revealed upregulation of proteins involved in transcription regulation, histone binding, protein-DNA complex assembly, amino acid biosynthesis, and protein translation machinery in the primary tumor and lung metastases. Proteins involved in extracellular matrix (ECM) assembly, cell-substrate adhesion, cholesterol transport, antioxidant activity, and complement activation pathways were downregulated in tumors (Suppl Fig. 1C). Protein abundance of the primary tumor was very similar to that of recurrent lung metastases, revealing similar dysregulation of cellular pathways compared to the adjacent normal tissues (Suppl Fig. 1B. Primary tumor and lung metastases showed indication of proliferative signatures, including upregulation of cell cycle pathways, DNA replication, transcription, translation and ribosome biogenesis (Fig. 1G). Conversely, ECM-receptor interaction, cell-cell adhesion, immune processes such as MHC protein complex and humoral immune response, complement activation, and antigen processing and presentation were downregulated in the primary tumor and all lung metastases (Fig. 1G).

### Increased one-carbon metabolism and serine addiction in SETTLE and orthogonal target validation

Metabolic reprogramming is one of the hallmarks of cancer, with particular significance in the altered one-carbon (1C) metabolism pathway (*29*). Among the differentially upregulated proteins, higher abundance (>2-fold difference) of serine biosynthesis proteins phosphoserine phosphatase (PSPH), phosphoserine aminotransferase 1 (PSAT1), and the one-carbon metabolism pathway protein serine hydroxymethyltransferase 2 (SHMT2) were observed in the primary tumor and all three lung relapses (Fig. 2A-D). Elevated levels of PSAT1 and PSPH have been identified as indicators of unfavorable prognosis in various types of tumors (*30–35*).

**Figure 2:**
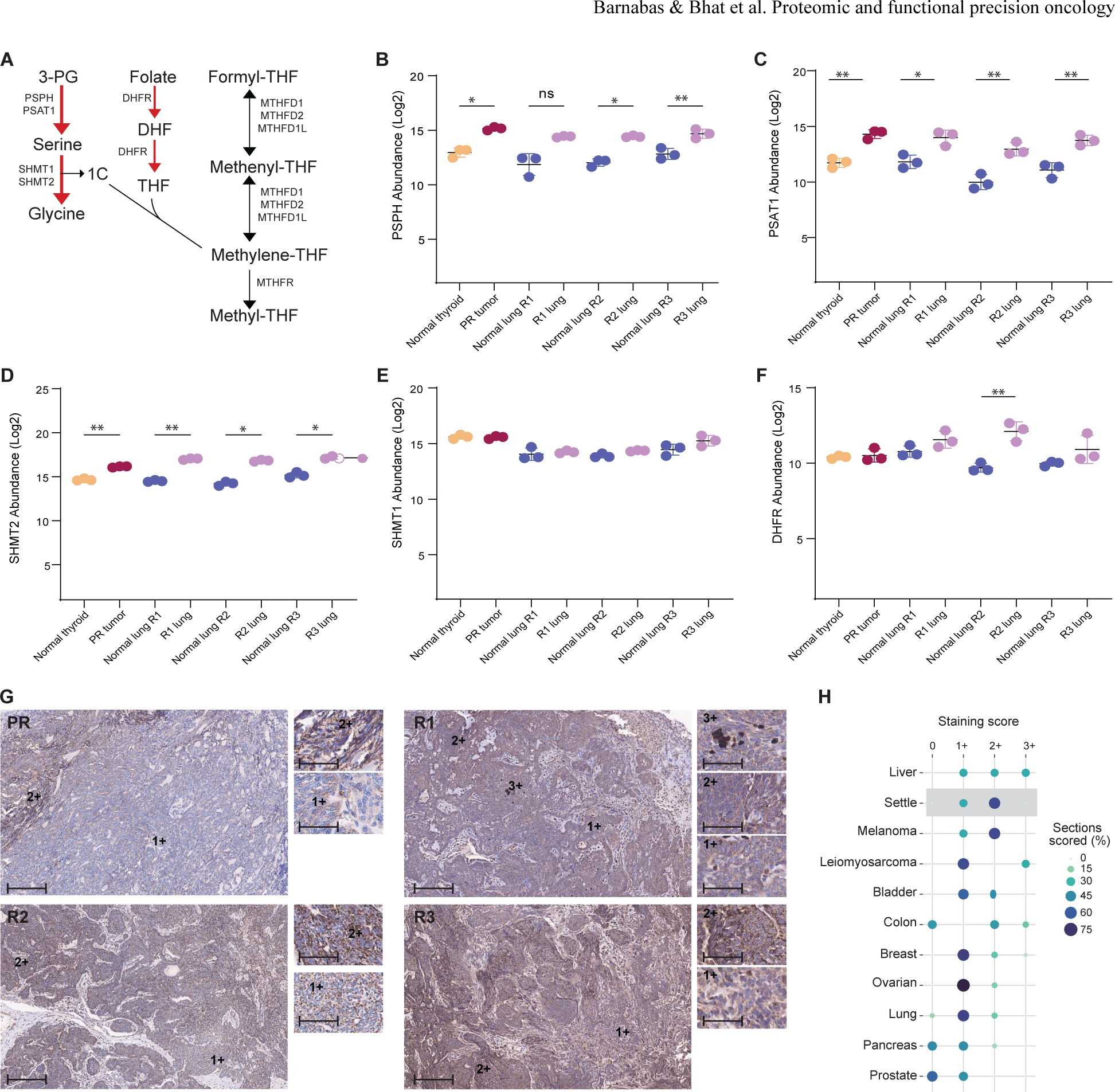
Increased one-carbon metabolism and serine addiction in SETTLE tumor. **A.** Schematic representation of serine biosynthesis and 1C metabolism pathways. Red arrows highlight the increased abundance of proteins involved in serine biosynthesis and those involved in serine and folate metabolism in the SETTLE tumor. **B-F.** Increased abundance of serine biosynthesis proteins PSPH (**B**), PSAT1 (**C**), 1C metabolism proteins SHMT2 **(D**), SHMT1 (**E**) and folate metabolism DHFR (**F**) in the primary tumor and lung metastases **G**. Validation of increased SHMT2 protein levels in SETTLE tumor using IHC staining. Adjacent normal thyroid and lung tissues show minimal basal level staining (score +1), tumor cells show more prominent staining (score +2), and granulocytes show the highest staining (score +3). Scale bars: 200 µm in main image; 60 µm in insets. **H**. Levels of SHMT2 in diverse tumor types using tumor microarrays; Liver (n=3), melanoma (n=3), and leiomyosarcoma (n=3) showed high SHMT2 levels; Bladder (n=7), colon (n=5), breast (n=14), ovarian (n=12) and lung (n=16) tumors showed moderate SHMT2 levels and pancreas (n=9) and prostate (n=7) tumors had the lowest SHMT2 levels. *p value <0.05 and **p value <0.01.

A close inspection of the 1C metabolism pathway revealed a subtle increase, albeit mostly statistically non-significant, in the abundance of many proteins, including SHMT1, dihydrofolate reductase (DHFR), methylenetetrahydrofolate dehydrogenase 1 (MTHFD1) and MTHFD2 (Fig 2E-F and Suppl Fig. 1D-E). SHMTs convert serine to glycine, thereby providing carbons for 1C metabolism to support *de novo* nucleotide and amino acid biosynthesis. MTHFD1 and MTHFD2 are integral components of the 1C metabolism pathway and exhibited increased abundance in one of the lung relapses compared to adjacent normal lung tissue (Suppl Fig. 1 D-E). DHFR, a key enzyme upstream in the 1C metabolism pathway, demonstrated elevated levels in the recurrent lung relapses, while no notable changes were observed in the primary tumor (Fig. 2F). DHFR, a well-established target to inhibit tumor growth, catalyzes the reduction of dihydrofolate to tetrahydrofolate and antifolate medications are recognized for their ability to inhibit DHFR. These findings showed that proteins in the 1C metabolism pathway were elevated in SETTLE tumor cells and suggested that inhibiting SHMT2 and DHFR could be an effective strategy to reduce tumor progression.

Pre-clinical studies in breast and lung cancer PDX indicate that SHMT2 constitutes an actionable target with the SHMT inhibitor sertraline (*36–39*). To obtain orthogonal validation of increased SHMT2 protein levels in SETTLE cells, we performed IHC staining for SHMT2. The primary tumor and the lung metastases showed positive IHC staining for SHMT2 in the tumor cell-enriched regions, whereas adjacent normal thyroid and lung tissues showed minimal staining (score +1), tumor cells with more prominent staining (score +2), and granulocytes with the highest staining (score +3) (Fig. 2G). We also evaluated spatial heterogeneity and found that SHMT2 was consistently elevated across the nodules. The turnaround time from biopsy to proteomic analysis and IHC validation was 15 days.

We then compared the SHMT2 levels in SETTLE tumor cells with those in tumors previously shown to be susceptible to SHMT2 inhibition in diverse pediatric and adult tumor types on tumor microarrays. By IHC analysis, SETTLE tumors manifested high SHMT2 levels similar to liver tumor, melanoma, and leiomyosarcoma, which had the highest number of tissue sections with increased staining (Fig. 2H and Suppl Fig. 1F). Bladder, colon, breast, ovarian and lung tumors showed moderate SHMT2 staining, and pancreas and prostate tumors had the lowest intensities (Fig. 2H and Suppl Fig. 1F). Breast tumors had been reported to be susceptible to SHMT2 inhibition (*37*), suggesting that the SHMT2 elevation found in SETTLE of this patient might render the malignant cells sensitive to SHMT2 targeted treatment.

### Primary tumor histology features and disease progression

The primary diagnostic (Dx) biopsy showed a spindle cell lesion with abrupt transitions to papillary-like structures and mature epithelial elements, including intestinal glandular epithelium, followed by a quick transformation into spindle cells. Noteworthy, the primary resection (PR, Fig. 1C) also demonstrated the biphasic morphology of dense non-descript spindle cells with an abrupt transition to intestinal-like epithelial glands. The spindled areas had a diverse morphology ranging from cell-dense areas, loose myxoid areas, and dense fibrotic bands. The Dx and PR have been previously reported (*25*).

The first metastatic specimen (R1, Fig. 1C) was a lung metastasis with a robust desmoplastic response separating the metastasis from surrounding normal lung parenchyma. The periphery of the metastasis showed a densely cellular pattern that predominantly consisted of mildly appearing spindle cells with prominent nucleoli. Delicate fibrovascular cores run through these cellular areas and the spindle cells became discohesive creating the papillary-like structures as seen in the primary resection. The second metastatic resection (R2, Fig. 1C) resulted in five metastasectomy specimens from the lung, all of which showed the peripheral desmoplastic response separating the tumor from normal lung parenchyma. All five specimens had fibrotic and densely cellular spindle cell components. Two pulmonary metastases had central fibrotic bands like the PR, and three had central sclerotic cores similar to the R1 specimen. Additionally, papillary-like structures were observed among the spindle cells with a myxoid extracellular component. The third metastatic resection (R3, Fig. 1C) resulted in two specimens that showed morphologic features consistent with the R1 specimen. Namely, a well-demarcated spindle cell population with dense spindle cell areas in the periphery and a more sclerotic central core. Immunohistochemistry for diagnostic purposes was done on all 3 relapses (R1, R2, R3), and the immunophenotype remained identical compared to the Dx biopsy and PR.

### Mapping tumor histology through SETTLE PDX models and SHMT2 expression analysis

To facilitate targeted drug analysis, viable cryopreserved specimens of the resected R2 were successfully grafted into a hind flank of NOD. Cg-Prkdcscid Il2rgtm1Wjl/SzJ mice (NSG-PDX) by subcutaneous injection and as onplants onto CAM (CAM-PDX). Histological investigation of NSG-PDX and CAM-PDX revealed morphological characteristics consistent with those seen in the patient-derived sample (Fig. 3).

**Figure 3:**
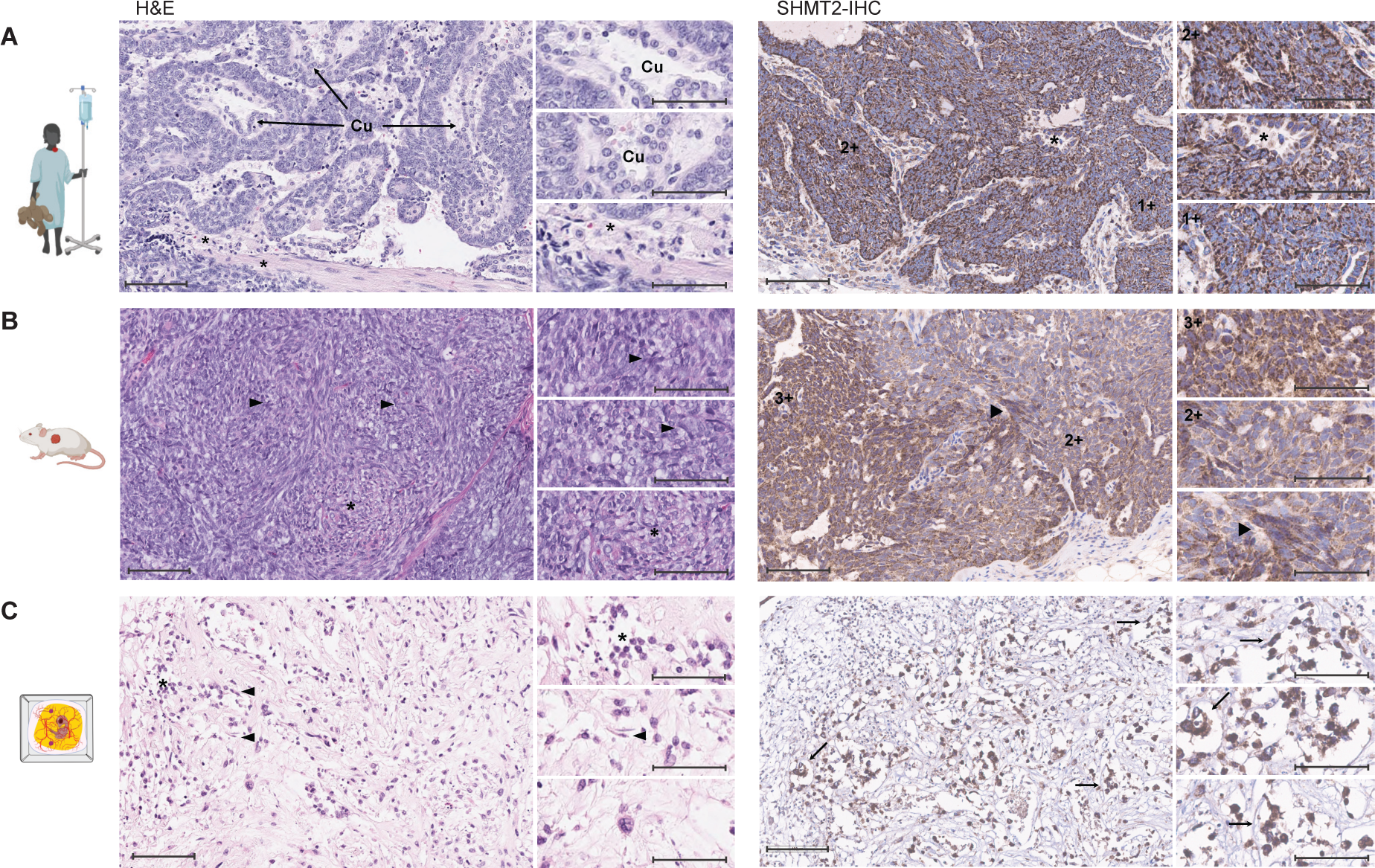
Retention of SETTLE tumor architecture in PDX models. **A.** Patient’s R2 sample. H&E staining (left) shows the pseudopapillary areas with a cellular thickness with a collagenous stroma (Arrowhead) that contains blood vessels and low-cuboidal lined epithelia (Cu). SHMT2 IHC (right) shows a diffuse 3+ and 2+ staining pattern in the tumor cells. The morphologic appearance of the SETTLE shows oval cells with spindled-like morphology. Some areas show a pseudo-papillary appearance with a central vascular core surrounded by layers of tumor cells. **B.** Mouse-PDX. H&E staining (left) shows a consistent pattern of cellular and spindle area as seen in the patient-derived specimen (black arrowheads). SHMT2 IHC (right) shows a heterogenous pattern, with a mix of 3+ and 2+ staining in tumor cells. **C**. CAM-PDX. H&E staining (left) shows largely discohesive cells with mild to moderate pleomorphism and hyperchromasia, tumor cells are oval-to-spindled in appearance with irregular nuclear membranes and scant cytoplasm. SHMT2 IHC (right) shows the retention of the staining seen in the primary human R2 and mouse-PDX samples. Scale bars: 100 µm in main image; 60 µm in insets.

The NSG-PDXs of primary SETTLE required 6-8 months to establish, with the resulting tumors achieving significant expansion and a cellular content of >80% human (Suppl Fig. 2). Histological examination of the NSG-PDX showed a dense collection of cells with unique round and spindle shapes, with a varied chromatin pattern characterized by irregular nuclear membranes and nucleoli (Fig. 3B). In addition, tightly packed cellular clusters resemble the patient surgical specimens, with cells exhibiting spindle-like features and forming structures resembling follicles, but no heterologous differentiation, like intestinal epithelium, was observed (Fig. 3A and B).

Histological examination of the CAM-PDX microtumors of SETTLE showed a distinct region of human cellular mass, encapsulated by CAM-derived epithelia within the Matrigel scaffold (Suppl Fig. 3). Atypical cells, either in clusters or as individual cells, are evident within the tumor mass. Tumor cells within the CAM-PDX displayed a predominance of discohesive growth pattern with mild to moderate pleomorphism and hyperchromasia, and often maintained a spindle or oval morphology (Fig. 3C). The nuclei often showed prominent nucleoli and irregular nuclear membranes, with occasional mitotic figures identified (Fig. 3A and C). SETTLE cells isolated from NSG-PDX tumors were also engrafted in larval zebrafish to establish zebrafish-PDX for additional pre-clinical assessment. The presence of SETTLE cells within the zebrafish-PDX were confirmed through H&E staining which highlighted morphological traits closely resembling the histological patterns of SETTLE tumors derived from mice, including high nuclear to cytoplasmic ratio, hyperchromasia and nuclear membrane irregularities (Suppl Fig. 4).

Immunohistochemical analysis of the corresponding CAM-PDX and NSG-PDX for SHMT2 showed a heterogenous pattern of expression resembling that seen in the patient R2, with a maximum score of 3+ and 2+ on the tumor tissue sections (Fig. 3A, B, C). In sum, the histopathological morphologies and expressions of SHMT2 of the established PDXs resemble those of the patient R2.

### Pre-clinical targeting of the 1C metabolism pathway as a metabolic vulnerability using personalized xenograft models

Identification of elevated SHMT2 protein abundance by global proteome profiling coupled with the ability to rapidly generate CAM-PDXs afforded an opportunity to rapidly evaluate targeting the 1C metabolic pathway. CAM-PDXs of R2 treated with SHMT1/2 inhibitors sertraline (Fig. 4A and B) or SHIN1 (Suppl Fig. 5) exhibited reduced tumoroid growth when compared to their respective vehicle controls. To corroborate this observation and determine a dose-response relationship, we used mouse cell-depleted dissociated SETTLE cells, expanded from NSG-PDX of R2, and found that these SETTLE cells were susceptible to sertraline at half maximal effective concentrations (IC50) between 5.5-10.1 µM (Fig. 4C).

**Figure 4:**
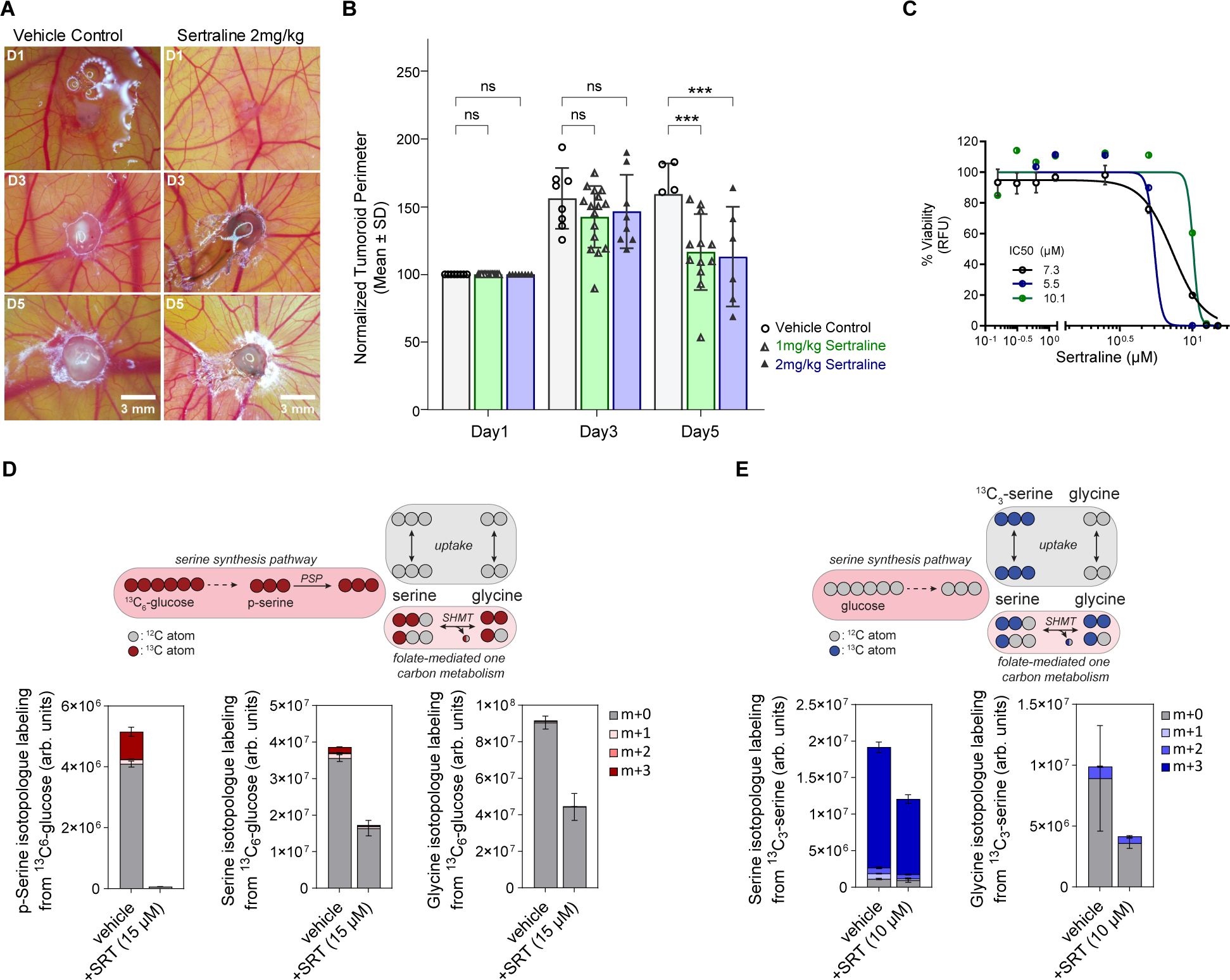
Pre-clinical targeting the 1C metabolic pathway in SETTLE. **A.** Representative images CAM-PDX of SETTLE tumor over five days with and without (Vehicle Control) treatment with sertraline. **B.** Measurements of CAM-PDX tumoroid perimeters normalized to Day 1 for untreated and treatment with 2 concentrations of sertraline (***P<0.001). **C.** In vitro viability assays of SETTLE PDX cells subjected to sertraline treatment (IC50 range 5-10 μM for 3 independently conducted assays). **D.** The abundance of p-serine, serine and glycine isotopologues from ^13^C-6-glucose labelled NSG-PDX cells treated with sertraline or the vehicle control. **E.** The abundance of serine and glycine isotopologues from ^13^C_3_-serine labeled NSG-PDX cells treated with sertraline or the vehicle control.

To validate that the observed response was indeed linked to the increased dependence on serine and glycine metabolism in SETTLE cells, we performed stable-isotope tracing using ^13^C_6_-glucose or ^13^C_3_-serine on NSG-PDX cells cultured *in vitro*. ^13^C_6_-glucose and ^13^C_3_-serine yield m+3 labeled serine through the serine synthesis pathway activity and uptake, respectively (Fig. 4D and E). Subsequent metabolism by folate-mediated 1C metabolism (FOCM), which includes SHMT1/2, leads to the shuffling of labeled and unlabeled carbons due to the cyclic nature of the pathway. Thus, the relative activity of SHMT1/2 can be inferred by quantifying the abundance of m+1 and m+2 versus m+3 serine isotopologues. Our results from a 12-hour culture with ^13^C_6_-glucose suggested that the activity of serine synthesis is relatively low in NSG-PDX cells with sertraline treatment, as m+3 serine reached only ∼5%. In contrast, cells cultured with ^13^C_3_-serine for 24 hours displayed ∼94% labeling split between m+1, m+2, and m+3 isotopologues and reduced abundance upon sertraline treatment. This suggested that SETTLE cells acquire serine primarily through environmental uptake. Further, the m+1 and m+2 isotopologues, which arise through FOCM flux, decreased by ∼20% with sertraline treatment.

### Sertraline and trimethoprim combination as a potential therapeutic approach for SETTLE tumors

There are additional druggable opportunities in the 1C metabolic pathway that may be exploited to elicit a more effective response (Fig. 5A). Cellular metabolic reprogramming is a hallmark of cancer progression and 1C metabolism involves folate and methionine progressions to generate 1C units for the biosynthesis of imperative anabolic precursors. 1C metabolism facilitates the biosynthesis of purine and thymidine production for the high needs of dividing cancer cells. It was shown that the antiproliferative effects of sertraline in combination with the mitochondrial inhibitor artemether led to cell-cycle arrest in the G1-S phase and antitumor activity against serine-dependent cell models of breast cancers (*37*). We found that combinations of sertraline and artemether exhibited drug synergy profiles in NSG-PDX cells comparable with those in the serine-dependent MDA-MB-468 cells, but not with the serine-independent MDA-MB-231 cells (Fig. 5B and Suppl Fig. 6). We also evaluated the combined treatment of sertraline with trimethoprim, an inhibitor of DHFR that was shown by proteomics to be significantly increased in SETTLE cells. *In vitro*, trimethoprim elicited little to no cytotoxicity on NSG-PDX cells as a single agent and insignificant synergistic effects in combination with sertraline (Fig. 5C). In contrast, both sertraline and trimethoprim inhibited the growth of SETTLE cells in zebrafish-PDX (established using cells from NSG-PDX) as single agents and elicited further inhibition in combination (Fig. 5D). To corroborate the *in vivo* results, CAM-PDX of SETTLE cells (also dissociated NSG-PDX) were treated with sertraline and/or trimethoprim (Fig. 5E); the results were suggestive of anti-tumor effects with sertraline and/or trimethoprim treatments.

**Figure 5:**
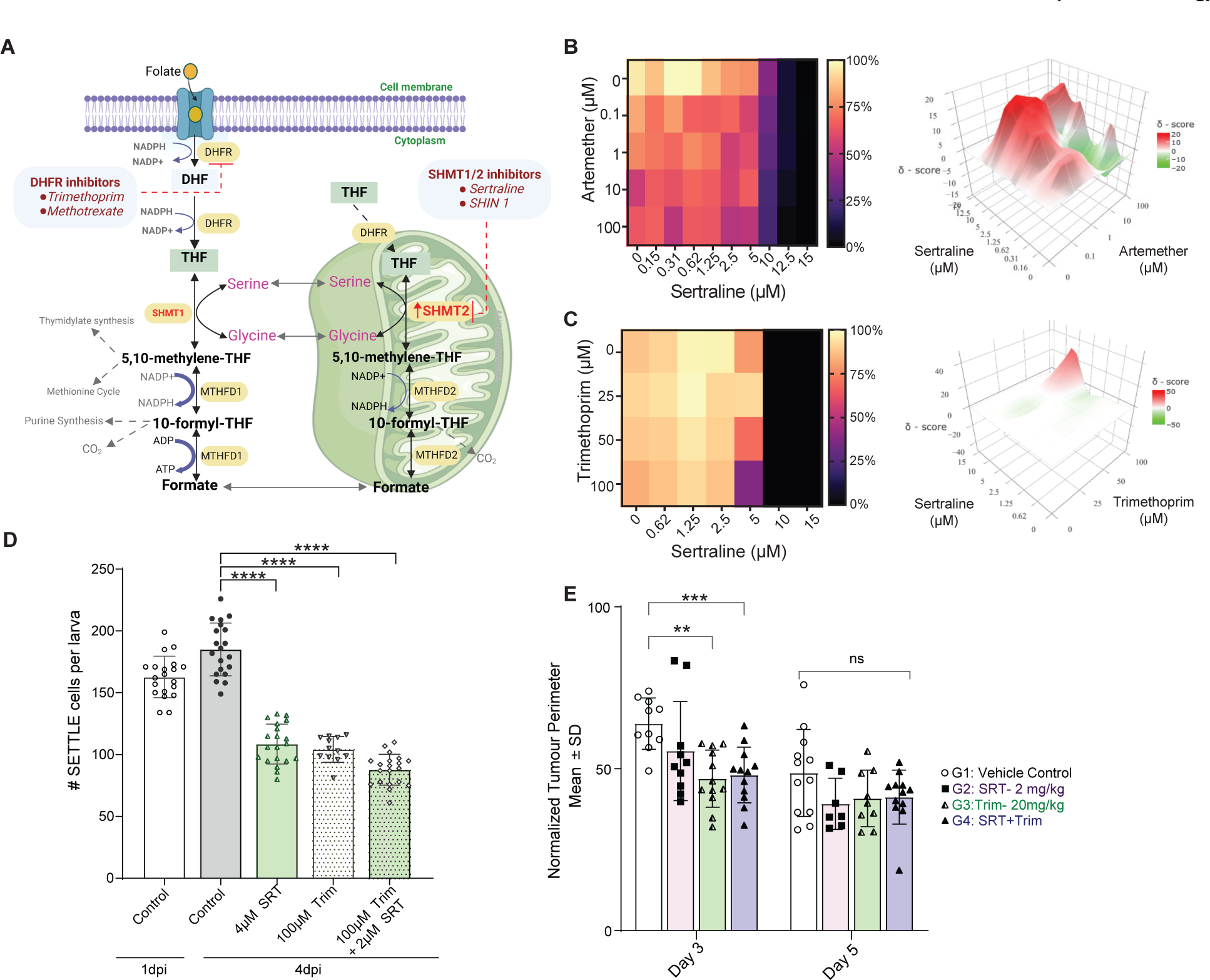
Sertraline and trimethoprim combination as a potential therapeutic approach for SETTLE. **A.** Schematic representation of the 1C metabolism pathway showing the significance of therapeutic inhibition of this pathway at DHFR and SHMT2 levels. Serine is converted to glycine by cytoplasmic SHMT1 (left) and mitochondrial SHMT2 (right). The 1C component sliced from serine is transferred to THF, generating methylene-THF. Therapeutic interventions, highlighted in red-dash lines (DHFR inhibitor: Trimethoprim, SHMT2 inhibitor: Sertraline), at two distant ends of this pathway stop the 1C unit being used for THF. THF is produced from folate and serves as a universal 1C acceptor. *DHF: dihydrofolate; THF: tetrahydrofolate; DHFR: dihydrofolate reductase; MFT: mitochondrial folate transporter; SHTMT1/2, serine hydroxymethyl transferase, cytosolic* (*1*)*/mitochondrial* (*2*)*; MTHFD1: methylenetetrahydrofolate dehydrogenase 1; MTHFD2: methylenetetrahydrofolate dehydrogenase 2 (2-like).* **B, C.** In vitro viability assays of SETTLE PDX cells subjected to treatment with sertraline and/or artemether (B) and sertraline and/or trimethoprim (C) shown as heat map viability plots (left) and the Highest Single Agent combinatorial drug synergy plots (d-score >10 indicative of synergistic effects) (right). **D.** SETTLE tumor cells engrafted in larval zebrafish were untreated or treated with sertraline (Sert) and/or trimethoprim (Trim). Scatter bar graphs depict the number of SETTLE cells per larva at 1- or 4 days post-implantation (dpi) (**** P< 0.0001). **E.** CAM engrafted with SETTLE tumor cells were untreated or treated with sertraline and/or trimethoprim. The scatter bar graph depicts the tumoroid perimeters at days 3 and 5 normalized against day 1. (***P<0.001).

### The patient treated with sertraline had a delayed and reduced tumor growth rate

The proteomic analysis and drug response from personalized xenograft models led a molecular tumor board to prioritize a three-month innovative therapy trial with sertraline for the patient. CT scans (Fig. 6A and B) encompassing the periods before initiation of sertraline therapy, during and after cessation of therapy revealed notable decreases in the monthly tumor growth rates at end of sertraline therapy, which further declined at 2 months post sertraline therapy, with no new parenchymal lesions forming (Fig. 6B, C and D). This suggests that the tumor responded to sertraline as a single-agent therapy, slowing the progression of tumor growth, although it did not reduce the tumor mass and RECIST scoring indicating progressive disease (Fig. 6A, D).

**Figure 6:**
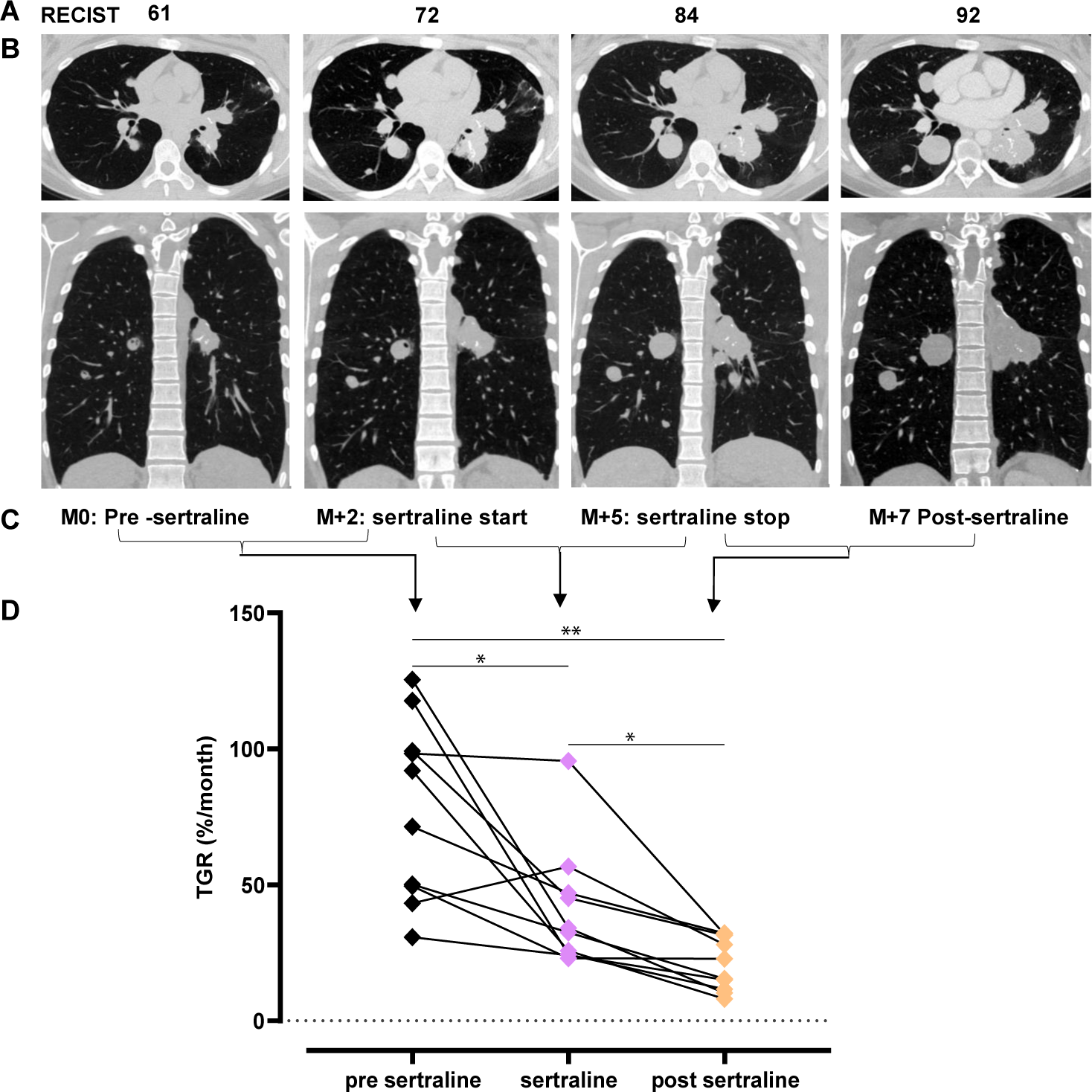
Tumor growth rate and RECIST score of SETTLE patient before and after sertraline therapy. **A-C.** RECIST score indicated in numbers (A), patient CT images before and after sertraline treatment (B) at 5-time points before, during, and after sertraline therapy spanning 7 months (C). **D.** graphs displaying monthly tumor growth rate tracked for 9 lesions. *p value <0.05 ; **p value <0.01.

## Discussion

We hypothesized that quantitative whole proteome profiling and drug response validation using patient-derived xenograft (PDX) models could provide on-time pre-clinical insight complementary to whole exome and transcriptome sequencing and inform clinical decision-making for pediatric precision oncology. To explore this in a real-world setting and establish a proof of principle, we identified a child diagnosed with a rare malignancy who had previously received genome profiling-guided innovative targeted therapy and was now experiencing progressive disease. Proteomics analysis identified SHMT2 elevation as an actionable target for sertraline therapy within two weeks and a positive response to sertraline was validated *in vitro* and in PDX models. During and following a 3-month innovative therapy trial with sertraline the patient showed progressive disease with reduced tumor growth rates.

Personalized precision oncology tailors the cancer treatment plan according to the unique molecular property of an individual’s tumor: it has the potential to transform cancer care by pinpointing the most effective treatment options, minimizing negative side effects, and improving patient outcomes. Despite its theoretical benefits, the success of precision oncology has so far fallen short of expectations, and applying this approach to complex and uncommon cancers in younger patients remains challenging and requires further research and development.

Precision medicine programs are enhancing multi-omics in-depth tumor profiling with functional testing of drug sensitivities in patient-derived models to support clinical decisions beyond existing approaches (*40*). Patient tissue xenografts preserve the original tumors’ characteristics, aiding in accurate representation. Rodent-based PDXs have been pivotal in establishing a wide range of models of pediatric solid tumors (*41*), with the caveat that these xenografts may require months to engraft. In comparison, CAM and larval zebrafish offer lower-cost and higher-throughput *in vivo* platforms for real-time pre-clinical validation of personalized therapies (*42, 43*). In this regard, therapeutic efficacy studies can be completed in CAM and larval zebrafish PDXs within a 2-week time frame. A prime limitation of all PDX platforms remains the availability of sufficient viable tumor tissues. Here, NSG-PDXs provided for the expansion of patient-derived material to facilitate further *in vitro* and *in vivo* testing even when primary tumor tissue has been exhausted.

Global quantitative proteome analysis identified elevated SHMT2 levels in the primary tumor, and subsequent lung relapses, as an actionable therapeutic target. Our findings of positive drug response to the SHMT2 inhibitor sertraline together with the DHFR inhibitor trimethoprim in PDX models suggest the potential of sertraline and trimethoprim combination therapy for SETTLE and other tumors dependent on serine/glycine synthesis, particularly when combined with drugs targeting key nodes of cancer cell metabolism. This observation contributes to the growing evidence supporting that proteome-based molecular profiling can identify actionable targets that are more reflective of the tumor phenotype and demonstrates the potential of proteomics and personalized xenograft models to provide timely pre-clinical support for medical decision-making in precision oncology.

This study demonstrates the ability of proteomics to provide actionable molecular insights beyond genomic alterations within days to weeks enabling its application in a personalized precision oncology context. We acknowledge that our study is limited to a single case making it impossible to assess how many and which patient population will benefit the most. To answer these important questions, we are now expanding this to children and adolescents with hard-to-treat cancers across Canada. However, we feel that it is important to report this first case immediately to widely encourage and support similar activities around the world. This study sets up the basis to implement proteomics into molecular tumor profiling of patients under PROFYLE and other pediatric precision medicine initiatives. Previous retrospective cohort studies have demonstrated that proteome analysis can identify drug targets and contribute to rational treatment choices in pediatric cancers (*28, 44*). Pediatric precision medicine initiatives such as ZERO and INFORM have showcased the application of functional precision oncology during diagnosis for various cancer types, thereby potentially offering more suitable treatment options. Furthermore, the integration of additional data layers, such as single-cell analysis, spatial profiling, and liquid biopsy models, including those considering proteoforms (*45*), leveraging the multiscale approach to enhance disease understanding and uncover novel biomarkers could be implemented (*46*). In addition, delving deeper into alternative approaches for combination studies could help identify the optimal fit or achieve better response outcomes, aiding in the prioritization of one combination over another in such scenarios. Therefore, these programs should engage patients at earlier stages and explore combination therapy strategies to improve the number of patients experiencing clinical benefits.

## Significance

This study emphasizes the benefits of integrating proteomics and personalized xenograft models to verify and complement therapeutic opportunities identified by genomics and expedite the evaluation of treatment responses for rare pediatric malignancies. The ability to provide timely, clinically relevant data invisible to established sequencing-focused precision oncology strategies offers a paradigm shift in medical decision-making and has the potential to significantly influence clinical practice.

## Acknowledgements

Data and/or materials used for this research were made available by the PRecision Oncology For Young peopLE (PROFYLE) program. PROFYLE has been supported by funds from many funders including Alberta Cancer Foundation, Alberta Children’s Hospital Foundation, BC Cancer Foundation, BC Children’s Hospital Foundation, Childhood Cancer Canada, Kids Cancer Care Foundation and Terry Fox Research Institute. This work was supported by the BC Children’s Hospital Foundation through the Better Responses through Avatars and Evidence (BRAvE) Initiative. Salary support was provided by the Michael Cuccione Foundation MCF (C.J.L., G.S.D.R., C.A.M., P.F.L., V.G.), the Canada Research Chairs Program (CRC-RS 950-230867, P.F.L.), the Canadian Institutes of Health Research (C.A.M., P.F.L.), the Michael Smith Foundation for Health Research Scholar Program (16442, P.F.L.), MITACS (T.A.B., G.B.) and the University of British Columbia (E.K.E.). Project support for J.A.C. and D.L.S. was provided by The Alberta Cancer Foundation. We thank all the authors for their contributions and technical assistance. This manuscript was edited at Life Science Editors. We are indebted to Dr Karla Williams (UBC Pharmaceutical Sciences) for vital assistance in establishing the CAM facility at BCCHRI. We gratefully acknowledge the participation of the patients and families that made this study possible and the BC Children’s Hospital staff physicians and Biobank staff for their tremendous efforts in collecting and maintaining specimens.

## Materials and Methods

### Ethical regulations

Patient specimens were collected by Biobank staff at BC Children’s Hospital. Consenting of the participant and collection of the specimen were conducted following review and approval by the University of British Columbia Children & Women’s Research Ethics Board (H17-01860, H18-02473-A037), and conformed with standards defined in the WMA Department of Helsinki and the Department of Health and Human Services Belmont Report. The studies using the chick embryo CAM model followed BCCHR institution internal protocols and University of British Columbia animal committee guidelines. Experiments with the CAM model in this paper adhered to Canadian Council of Animal Care (CCAC) regulations that did not require approval and ARRIVE guidelines 2.0 (PMID: 32663219). Additionally, avian embryos are not considered live vertebrate animals under NIH PHS policy. The use of zebrafish in this study was approved and carried out according to the policies of the University of Ottawa’s Animal Care Committee (Protocol #CHEOe-3195), which is governed and certified by the Canadian Council on Animal Care. All mouse procedures and experiments were reviewed and approved by the University of Calgary Animal Care Committee (AC21-0016).

### Tissue processing

Clinical tissues used in this study were received fresh from the operating room within 30 minutes of removal. Tissues were triaged into either, snap frozen samples using liquid nitrogen cooling or viably cryopreserved using 10% DMSO with fetal bovine serum (FBS). The samples were processed into blocks using 10% neutral-buffered formalin and paraffin embedding. Slides were sectioned at 4 microns for hematoxylin and eosin (H&E) staining. CAM-generated tumoroid tissues were removed, quickly washed in DPBS, fixed in 10% formalin for 24-48 hours, and processed for paraffin embedding (FFPE).

### Immunohistochemistry (IHC)

Human SETTLE tissue, Mouse PDX, CAM-PDX SETTLE, and tissue microarray blocks were sectioned at 4 & 5 µm using Thermo Scientific Rotary Microtome HM340E. The slides were dried at 37°C overnight and briefly baked at 65°C before sections were deparaffinized in xylene, then rehydration steps using an ethanol gradient of 100%, 95%, 75%, and 50%, followed by a final wash in double distilled water. Hematoxylin and Eosin (H&E) staining and Immunohistochemistry (IHC) were performed as per the manufacturer’s instruction (Vector Laboratories, USA). Briefly, a steam-generated heat-induced epitope retrieval (HIER) method was used for antigen retrieval by citrate buffer using a steam cooker with antigen retrieval solution (10 mM Na citrate pH 6 + 0.05% Tween 20). Deparaffinized and hydrated sections were incubated for 10 min at room temperature with 3% H2O2 to block endogenous peroxidase activity. After a rinse with PBS, the slides were blocked for unspecific binding with protein Avidin solution (goat serum in PBS) for 20 mins, at room temperature. Anti-SHMT2, a rabbit polyclonal antibody (Proteintech, USA; Cat No. 11099-1-AP) was added at a concentration of 1 µg/ml and incubated overnight at 4°C. The biotinylated secondary antibody was added to a blocking buffer and incubated for 30 min at room temperature. Slides were washed with PBS and then the Horseradish peroxidase (HRP) enzyme conjugated to avidin (ABC reagent) was added for 30 min at room temperature. Similarly, the LAMP1, an anti-human CD107a antibody (clone H4A3, Biolegend, USA) was applied at the concentration of 0.5 mg/mL and incubated overnight at 4°C, the procedure was followed by PBS washings and by the addition of biotinylated secondary antibody (HRP) streptavidin conjugate (1:200 for 30 min) and finally incubated with ABC reagent for 25-30 min, (VECTASTAIN Elite, Vector Laboratories, USA). An isotype control IgG antibody was maintained against both the primary antibodies in parallel. Slides were washed with PBS and incubated with Liquid DAB+ (3-3’-Diaminobenzidine) substrate-chromogen System (Dako, North America) according to the manufacturer’s protocol. After a rinse in water, the slides were counterstained with Hematoxylin, dehydrated, cleared, and mounted with coverslips by ShurMount and Cytoseal 60 (Epredia). All H&E and IHC slides were scanned with a Leica Aperio AT2 slide scanner and images were processed with Aperio ImageScope (Leica biosystems v.12). Tumor orientation was maintained consistently during the study to observe growth and dissemination during the implantation period.

### Whole genome and transcriptome sequencing, and data analysis

Tumor and normal genomes were sequenced on HiSeqX using v2.5 chemistry. Transcriptomes were sequenced on NextSeq500 using v2 chemistry. Normal and tumor reads were aligned to the human reference genome (hg38) using the Burrows-Wheeler Alignment miniMap2 tool (v2.15). Somatic point mutations (SNVs) and small insertions and deletions (indels) were detected using Strelka (v2.9.10) and Mutect2 (v2.4.0). Somatic copy number alterations were identified using CNAseq (v0.0.6), and loss of heterozygosity used APOLLOH (v0.1.1). Structural variants (SVs) in RNA-Seq data were identified using the assembly-based tools ABySS (v1.3.4) and TransABySS (v1.4.10) and alignment-based tools Chimerascan (v0.4.5) and DeFuse10 (v0.6.2); SVs in the DNA sequence data were identified using assembly-based tools ABySS and Trans ABySS and alignment-based tools Manta v1.0.0 and Delly v0.7.3. Putative SV calls identified from the DNA and RNA sequences were merged into a consensus caller MAVIS (v2.1.1).

RNA-seq reads were aligned using STAR (v.2.5.2b) to the human reference (hg38) and expression was quantified using RSEM (v.1.3.0). Publicly available transcriptome sequencing data from Illumina BodyMap 2.0, the Genotype-Tissue Expression (GTEx) Project, The Cancer Genome Atlas, Treehouse Childhood Cancer Initiative, and the TARGET program were used to explore the expression profiles of human genes and transcripts.

### Automated sonication-free acid-assisted proteome (ASAP) tissue lysis workflow

Primary pretreatment resection (Primary), first (R1), second (R2) and third (R3) relapse were utilized for the proteomics analysis. FFPE specimens were fixed in 10% neutral-buffered formalin, 10 µm sections from FFPE blocks were mounted on positively charged glass slides and dried at 37°C overnight. After deparaffinization and H&E staining, the slides were macrodissected into the tumor and adjacent normal tissue by a pathologist. Each timepoint was processed in triplicate, where each triplicate consisted of three 10 µm thick tissue sections. The macro dissected H&E-stained tissues were processed as described in Barnabas et al. (PMID 36724070). Briefly, samples were resuspended in 25 µl 75% trifluoroacetic acid (TFA) for 10 min at room temperature. 175 µl Neutralization buffer (2 M Tris, 3% SDS) was added and incubated at 95°C for 80 min followed by reduction and alkylation with 10 mM TCEP and 40 mM CAA at 95°C for 20 min.

Proteins were extracted by SP3 as published (PMID 30464214) using the Thermo KingFisher Apex platform. 10 µl hydrophobic SpeedBead Magnetic Carboxylate bead (stock solution 50 mg/ml; G&E 651521050250) and 10 µl hydrophilic SpeedBead Magnetic Carboxylate bead (stock solution 50 mg/ml; G&E 451521050250) were mixed per sample and bound to a magnet for 1 min. The beads were washed 3 times with 1 ml of MS-grade H2O and protein binding was induced by adding a final concentration of 80% Ethanol for 10 min. The SP3 beads were washed three times with 90% Ethanol followed by on-bead protein digestion in 80 µl 100 mM Ammonium bicarbonate containing 0.5 µg Trypsin/LysC Mix (Promega V507A) (enzyme/protein ratio of 1:50). The digestion mixture was incubated for 16-18 hrs at 37°C. Peptides were eluted from the SP3 beads using the KingFisher Apex robot and the peptide concentration was measured using the Pierce Quantitative Colorimetric Peptide assay according to the manufacturer’s protocol. Peptides were purified on Empore SPE C18 STAGE tips (PMID 17703201) or BioPureSPN PROTO 300 C18 Mini or Midi columns (The Nest Group Inc.) according to the manufacturer’s protocol. Purified peptides were concentrated using a vacuum concentrator and resuspended in 0.1% Formic Acid (FA) containing iRT peptides at 1:300 final dilution (Biognosys).

### Liquid chromatography and mass spectrometry

1 µg peptide per sample were injected into an Easy-nLC 1200 liquid chromatography system coupled to a Q-Exactive HF Orbitrap mass spectrometer (Thermo Scientific). Buffer A contained 2% acetonitrile (ACN) and 0.1% FA and Buffer B was 95% ACN and 0.1% FA. The peptides were separated on a 50 cm µPAC nano-LC column (reverse phase C18, pore size 100-200 Å, Thermo Scientific) with a flow rate of 300 nL/min and a gradient of 2 to 26% buffer B over a 180-minute gradient. A full MS scan with a mass range of 300–1650 m/z was collected with a resolution of 120,000. Maximum injection time was 60 ms and AGC target value was 3×106. DIA segment MS/MS spectra were acquired with a 24-variable window format with a resolution of 30,000. AGC’s target value was 3×106. Maximum injection time was set to ‘auto’. The stepped collision energy was set to 25.5, 27.0, and 30.0.

De-salted peptides from samples were combined into a single pool and analyzed in ten gas-phase fractions by injecting 1μg per in a 3 h gradient. The first eight fractions (340–820 m/z) were analyzed over a 60 m/z window (e.g., fraction 1: 340–400 m/z), each with a loop count of 30 and a window size of 2 m/z. The last two fractions (820–1180 m/z) were analyzed over a 180 m/z window each, with a loop count of 30 and 6 m/z window.

Proteomic raw data was searched using Spectronaut Pulsar X (Biognosys) using a human FASTA from UniProt (Dec 2021) and a FASTA file for iRT peptides (Biognosys) was included in the search. For the search, enzyme, and digestion type were set to Specific, and Trypsin/P, acetyl (Protein N-term), and oxidation (M) were set as variable modifications and carbamidomethyl (C) was set as fixed modification. Maximum and minimum peptide length were set to 7 and 52 amino acids, respectively and missed cleavage was set to 2. Precursor and protein FDR were set to 1% and a minimum of 2 peptides were used for quantification. The data was normalized in Spectronaut based on precursors identified in 70% of the samples. Data was filtered for proteins identified in 70% of tumors or adjacent normals and missing values were imputed using a down-shifted normal imputation strategy in perseus (PMID 27348712). Student’s t-test was performed using perseus and proteins with log2 fold change >1 with an FDR threshold of 5% was filtered for enrichment analysis. Gene ontology enrichment analysis was performed using GOrilla (PMID: 19192299). We utilized the pyCirclize package from https://github.com/moshi4/pyCirclize for the circular plot. The log2 fold-change values, calculated from tumor vs normal comparison, are displayed on the proteomics side of the plot. The genes depicted are those targeted by at least one of the commonly used drugs in pediatric cancer. For the dotplot, cancer versus normal testing is performed using independent sample t-tests with Benjamini-Hochberg correction. The p-value is as follows: not significant: p <= 1.00e+00, *: 1.00e-02 < p <= 5.00e-02, ** : 1.00e-03 < p <= 1.00e-02, *** : 1.00e-04 < p <= 1.00e-03, **** : p <= 1.00e-04

### Establishment of SETTLE PDXs in NSG mice

Fresh tumor tissue samples were collected at the time of surgery by the PROFYLE West biobank at British Columbia Children’s Hospital (BCCH) and preserved in vapor phase liquid nitrogen. A portion of the cryopreserved human tumor sample was used for the establishment of a patient-derived murine xenograft, under institutional review board (IRB) approval (HREBA.CC-16-0144). Six-to eight-week-old female NOD.Cg-Prkdcscid Il2rgtm1Wjl/SzJ (NSG) mice (Jackson Laboratories, Bar Harbor, MA, USA) were housed in groups of five and maintained as the procedures reviewed and approved by the University of Calgary Animal Care Committee (AC21-0016). Briefly, NSG mice were anesthetized via intraperitoneal injection of 50 mg/kg of ketamine and 5 mg/kg of xylazine. A 5 mm section of tumor tissue was implanted subcutaneously through an incision on the right hind flank. Following surgery, mice were administered two doses of buprenorphine (0.05mg/kg, subcutaneously), 12 hours apart, and monitored three times per week for signs of tumor growth. Once tumors had established (6-8 months), the mouse was sacrificed (intraperitoneal injection of 100 mg/kg ketamine and 10 mg/kg xylazine followed by cervical dislocation), and the tumor was collected (*in vivo* passage 1). Tumor tissue was then implanted into 5 mice (as described above) collected and stored. The presence of human tumor cells in PDX-generated samples was confirmed by immunohistochemistry using human-specific nucleolin antibody (Supplemental Fig. 3).

To prepare dissociated SETTLE PDX tumor cells, surgically removed NSG-PDX tissue was rinsed in sterile saline and placed on ice. Tissue was cut into 5 mm pieces and enzymatically dissociated using the Miltenyi Human Tumor Dissociation Kit and the Miltenyi GentleMACS Octo (with heaters) dissociator system (program: 37C_h_TDK_2). Following dissociation, the sample was filtered through a 70 µm filter, cells were counted, centrifuged at 300 x g for 7 minutes, supernatant discarded, and the cell pellet subjected to the Miltenyi Mouse Cell Depletion Kit (Miltenyi Biotec). The purified human cells were then aliquoted into 2 mL cryovials at a density of 3–5 million cells in 1 mL of Cryostore CS10 and stored as described above.

### Establishment of SETTLE PDXs in CAM

Fertilized eggs from White Leghorn chickens (Gallus gallus domesticus) were purchased from the University of Alberta, Edmonton, Alberta. Following overnight reset of the air sac at 14°C, eggs were placed into a hatcher incubator (Digital 1502 sportsman, GQF, Berryhill, ON Canada) at 37°C with relative humidity >70% for 4 days. The *ex-ovo* method for tumor engraftment was used for all CAM-PDX studies. Eggs were cracked aided with a rotary cutting tool (Dremel) and the deshelled chorioallantoic membrane (CAM) was placed in lidded sterile weigh boats (Cat. No. 08-732-113, Fisher Scientific). CAMs were further incubated at 37°C with humidity >70% until embryonic day 11 (ED11), when tumors will be onplanted.

Viably cryopreserved primary SETTLE tumors were thawed rapidly, the tissues rinsed with Dulbecco’s phosphate-buffered saline (DPBS, Gibco, Waltham, MA) and submerged in fetal bovine serum (FBS, Gibco, Waltham, MA). Tumor fragments were cut into smaller 1-2 mm pieces for implantation. Tumor fragments were onplanted onto gently lacerated membrane of ED11 CAMs, and immediately overlayed with 15-20 µL of cold Geltrex (Lot#A1413.02 & A14132-02, Gibco, NY USA). Tumor onplanted CAMs (herein referred to as tumoroids) were incubated at 37°C with humidity for up to 4-7 days, depending on the experimental workflow with routine imaging conducted to assess tumoroid growth. Tumors were typically harvested between D4-D7 following implantation for cryopreservation in 10%DMSO with RPMI 1640 media (Gibco, Waltham, MA), or formalin-fixed for paraffin embedding (FFPE) and histological workups.

### *In vitro* targeted drug response of SETTLE PDX cells

Targeted drug response was assessed *in vitro* using the aforementioned dissociated and cryopreserved SETTLE PDX cells (from mouse depleted PDX tumors). Cells were thawed rapidly in a 37°C water bath, transferred to 10mL CoDMEM (DMEM without pyruvate (Corning), with 10% FBS (Invitrogen) and penicillin-streptomycin (Invitrogen)), spun down, and resuspended either in fresh CoDMEM with 10% FBS or 10% dialyzed FBS. Where indicated, SETTLE cells were cultured in standard tissue culture dishes for up to 7 days in CoDMEM prior to drug assays. For comparative purposes, some experiments utilized MDA-MB-231 and MDA-MB-468 cell lines (ATCC HTB-26 and HTB-132) maintained at 37°C, 5% CO_2_ in cDMEM (DMEM (Sigma-Aldrich), with 10% fetal bovine serum (FBS, Invitrogen), penicillin-streptomycin (Invitrogen), and non-essential amino acids (Invitrogen)), and switched to CoDMEM for the drug assays.

Cells were lifted using trypsin-EDTA, washed and resuspended in media. 5000 live cells were plated per well of a 96-well plate and allowed to adhere overnight. Next day, the media was drained and replaced with drug (sertraline, arthemeter and/or trimethoprim) containing media (pre-diluted at the indicated concentrations) and incubated for 4 days. Cell viability was assessed with CellTiter-Glo Luminescent Cell Viability Assay (Promega), according to manufacturer’s instructions. Prism (GraphPad) was used to generate dose response plots and to calculate IC50 values and statistics. Additional plots and calculations of combinatorial drug synergies using the HSA (Highest Single Agent) reference model utilized SynergyFinder3.0 (PMID: 35580060).

### Targeted drug response of SETTLE in CAM-PDX

Prior to drug treatments in CAM-PDXs, preliminary safety studies were conducted for each drug to assess the maximum tolerated dose in non-tumor-bearing CAMs. For primary SETTLE CAM-PDXs, sertraline (S4053, Selleckchem, USA), SHIN1 (#6998, Tocris), or vehicle control were administered directly onto the tumoroids, on the day of implantation (D1) admixed with Geltrex, and with topical redosing at D3. Treatment response was assessed by imaging the CAM-PDX tumoroids at regular intervals (D1, D3, D5) (Motic SMZ-171 stereo microscope equipped with an Olympus DP72 color camera and cellSens Standard (Olympus Corp., Tokyo, Japan). The tumoroid periphery/area was measured using ImageJ software.

A modified protocol was used to assess treatment response to sertraline and/or trimethoprim of mouse-expanded SETTLE tumors in CAM-PDXs (primary non-expanded tumors had been exhausted). Viably cryopreserved mouse PDX tissue pieces were rapidly thawed, washed in DPBS, and processed by scalpel-based cutting into numerous small pieces. The tumor pieces were then pressed gently with a syringe plunger (#309602, BD) to dissociate the tissue into a homogeneous cell suspension that was passed through a cell strainer (40µm, CT#22363547, Fisher brand). The cells were pelleted by centrifugation, and the cell pellet resuspended in RPMI media in sufficient volume for the requisite group numbers in the study. To prepare gel spheroids for implantation, the tumor cell suspension was mixed with an equal volume of Geltrex and pipetted onto a pre-warmed petri dish using the hanging drop method to form dome-shaped spheroids. After allowing the spheroids to set at 37°C for 30 mins, complete media was added and further incubated for 2 hours until onplantation on lacerated CAMs. Tumoroids received topically administered treatments of vehicle only control, sertraline and/or trimethoprim at the indicated concentrations at the onset of implantation (D1) and re-applied on day 3 (D3).

### Drug treatment response of mouse-derived cells in larval zebrafish PDX

Adult *Casper* zebrafish (PMID 18371439) were bred in accordance with standard protocols (Westerfield et al., 2000) and maintained under controlled conditions (28°C, 14h:10h light: dark cycle) in the aquatics facility at the University of Ottawa. All experimental procedures were conducted in compliance with the University of Ottawa’s Animal Care Committee policies (Protocol #CHEOe-3195), which adhere to the guidelines of the Canadian Council on Animal Care. The study was conducted in compliance with animal care committee policies and guidelines. To assess the toxicity of drugs of interest, in vivo studies were conducted on Casper zebrafish larvae using immersion therapy. Cryopreserved dissociated SETTLE cells (mouse expanded) were thawed and processed using a dead cell removal kit (Miltenyi Biotec, #130-090-101) to obtain viable single cells, which were then labeled with CellTracker DeepRed cytoplasmic fluorescent dye (ThermoFisher, #C34565). The labeled cells were resuspended in Dulbecco’s Modified Eagle Medium (DMEM) supplemented with 10% heat-inactivated fetal bovine serum (FBS) for injection into 48-hour post-fertilization (hpf) zebrafish larvae. Larvae were anesthetized with tricaine (0.3mg/mL) and arrayed in agarose injection plates for cell transplantation. Using a pulled capillary needle, 100-200 cells were manually injected into the yolk sac (YS) of each larva, followed by incubation at 35°C. One day post-injection (1 dpi), larvae were screened under a Far-Red filter (620-700 nm) to confirm the presence of human tumor cells in the YS. Larvae with tumor engraftment were divided into groups and treated with solvent control, single agents, or drug combinations via immersion therapy for 72 hours (about 3 days). To evaluate drug response, *ex vivo* tumor cell quantification was performed at 1 dpi (baseline/untreated larvae) and 4 dpi (endpoint). Larvae were dissociated in a collagenase solution, and the resulting cell suspension was centrifuged and washed before resuspending in 10 µL per embryo for imaging. Post-quantification, fluorescent tumor cell numbers were assessed using FIJI software.

### Data analysis

All CAM experimental data were analyzed using GraphPad Prism10.0 software. Results were expressed as the Mean ± SD Comparisons between treatments were performed using the either two-way or one-way ANOVA test including the Tukey post-hoc test. Values represented as non-significant (ns) ≥ 0.05, * < 0.05, ** < 0.01, *** < 0.001, **** < 0.0001. Larval zebrafish PDX data sets were analyzed (GraphPad PRISM 9.0 software) by using one-way ANOVA with Dunnett’s multiple comparison test. Differences were considered significant when P was < 0.05 and statistical output was represented as follows: non-significant (ns) ≥ 0.05, * < 0.05, ** < 0.01, *** < 0.001, **** < 0.0001.

### Metabolic tracing

A vial of cryopreserved SETTLE PDX cells (from mouse depleted PDX tumors) was quickly thawed, resuspended in pre-warmed stable-isotope tracing media, centrifuged at 300xg for 5 minutes to remove cryopreservation media, resuspended in tracing media, and plated equally onto 6-well plates containing either vehicle (DMSO) or 10 µM sertraline. Stable-isotope tracing media was formulated by supplementing custom DMEM lacking glucose and amino acids (US Biological, D9800-27) with 10% dialyzed fetal bovine serum and ^13^C_6_-glucose/unlabeled glucose, ^13^C_3_-serine/unlabeled serine, and other unlabeled amino acids; as indicated. Following the indicated culture period, cells were quenched by quickly aspirating media and washing with ice cold 0.9% NaCl, prepared in HPLC-grade water (Sigma, 270733), before adding 1 mL of ice-cold extraction buffer. The extraction buffer consisted of 80% methanol (VWR, BDH20864.400), 20% water (Sigma, 270733). Cells were scraped into extraction buffer, transferred to tubes, vortexed at 4°C for 5-10 minutes, and centrifuged at 4°C at maximum speed for 15 minutes. 0.9 mL of supernatant was transferred to a new tube and concentrated using a SpeedVac (Thermo Fisher, SPD120) until dry. Metabolites were reconstituted into 25-50 µl of water, vortexed, centrifuged, and transferred to vials for analysis by LCMS.

Samples were loaded into a temperature controlled (6°C) autosampler and subjected to an LCMS analysis to detect and quantify peaks corresponding to annotated metabolites. A ZIC-pHILIC (2.1 x 150 mm, 5 µm; Millipore) LC column was coupled to a Vanquish LC (Thermo Scientific). The volume oven temperature was set to 25°C, maintained by forced air and an integrated column heater. The column was pre-equilibrated using a flow rate of 100 µl/min and 80-20%B. Following injection of 2 µl of sample, the following gradient elution at 100 µl/min was used: 80-20%B (0-30 min), 20-20%B (30-40 minute), and 20-80%B (40-40.5 minute); the LC column was re-equilibrated using 80-80%B from 40.5-52 minute before subsequent injections. Mobile phase composition was: (A) 10 mM ammonium carbonate in HPLC-grade water, pH 9.0 and (B) acetonitrile, 100%. Mobile phase A was freshly prepared or used within one week.

The LC was coupled to an Exploris 240 (Thermo Scientific) mass spectrometer operating in heated electrospray ionization mode (HESI) for analysis. The following parameters were set for HESI: spray voltage 3.4 kV (positive) and 2 kV (negative), static spray voltage, sheath gas 25, aux gas 5, sweep gas 0.5, ion transfer tube temperature 320°C, and vaporizer temperature 75°C. The global parameters included an expected peak width of 20 seconds, mild trapping, and a default charge state of 1. A 40-min polarity switching data-dependent Top 5 method was used for positive mode and a data-dependent Top 3 method was used for negative mode. Full MS scan parameters for both positive and negative modes were set as follows: scan range 67-1000 *m/z* collected in profile mode, Orbitrap resolution 120,000, RF lens 70%, AGC target of 300%, and maximum injection time set to automatic. ddMS2 for positive mode were collected in centroid mode at an Orbitrap resolution of 30,000, isolation window of 1.5 ^m/z^, an AGC target set to standard, a maximum injection time set to automatic, and a normalized collision energy set to 10%, 30%, and 80%. ddMS2 for negative mode were collected in centroid mode at an Orbitrap resolution of 30,000, isolation window of 2 *m/z*, an AGC target set to standard, a maximum injection time set to automatic, and a normalized collision energy set to 30%. For both positive and negative ddMS2, we applied an intensity threshold of 5e4 and a dynamic exclusion of 5 ppm for 10 seconds, excluding isotopes.

Raw LCMS data were analyzed using TraceFinder (versions 5.1 and 5.2, Thermo Scientific). Peaks corresponding to specific metabolites were annotated using a retention time and accurate mass library created from the Mass Spectrometry Metabolite Library of Standards (MSMLS, IROA Technologies) and other authentic standards acquired from Sigma. Peak areas were used to quantify the relative abundance of each metabolite and were quantified using the following general ICIS algorithm settings: a m/z discrimination threshold of 5 ppm, a retention time window of 60 or 120 seconds, a minimum peak area of 5e5, a smoothing factor of 9, an area noise factor of 5, a peak noise factor of 10, a baseline window of 100, a minimum peak height signal-to-noise ratio of 2, a minimum peak width of 5, a multiplet resolution of 10, an area tail extension of 20, and an area scan window of 0. Each peak integration was manually inspected, and individual settings were adjusted to optimize integration.

### Data availability

To maintain the patient’s and family’s privacy in this study of a single case the raw data is not made available at this point. Release of the raw data as part of an aggregated patient cohort is in preparation. Reviewers may request access to the data stored on secure systems.

## Supplementary Material

**Suppl Table 1:**
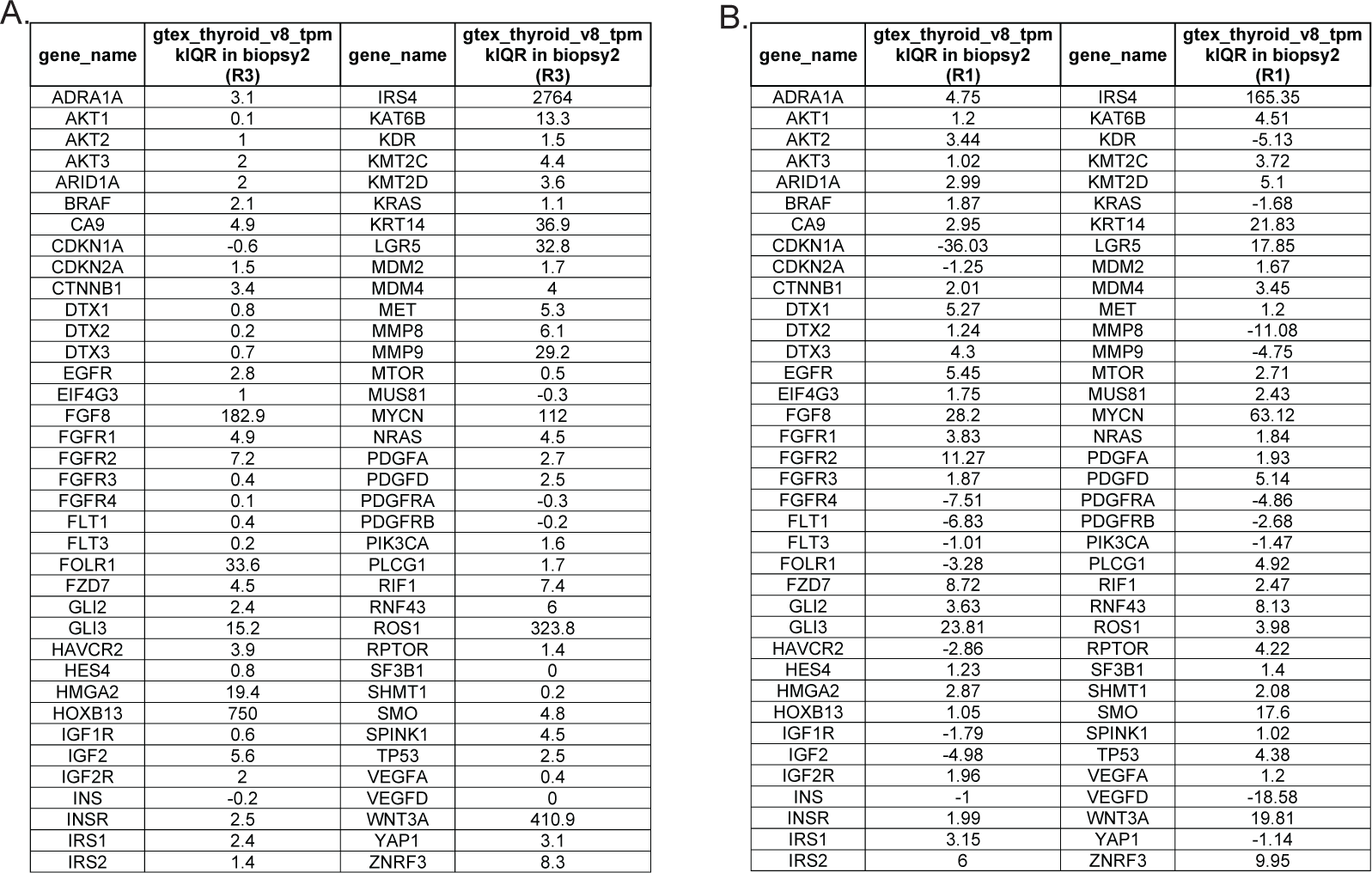
Transcriptome analysis of SETTLE relapses. Whole genome analysis of the thoracoscopic biopsy of (**A**) relapse R3 and (**B**) relapse R1. Gene expression is compared to normal thyroid tissue.

**Suppl Figure 1:**
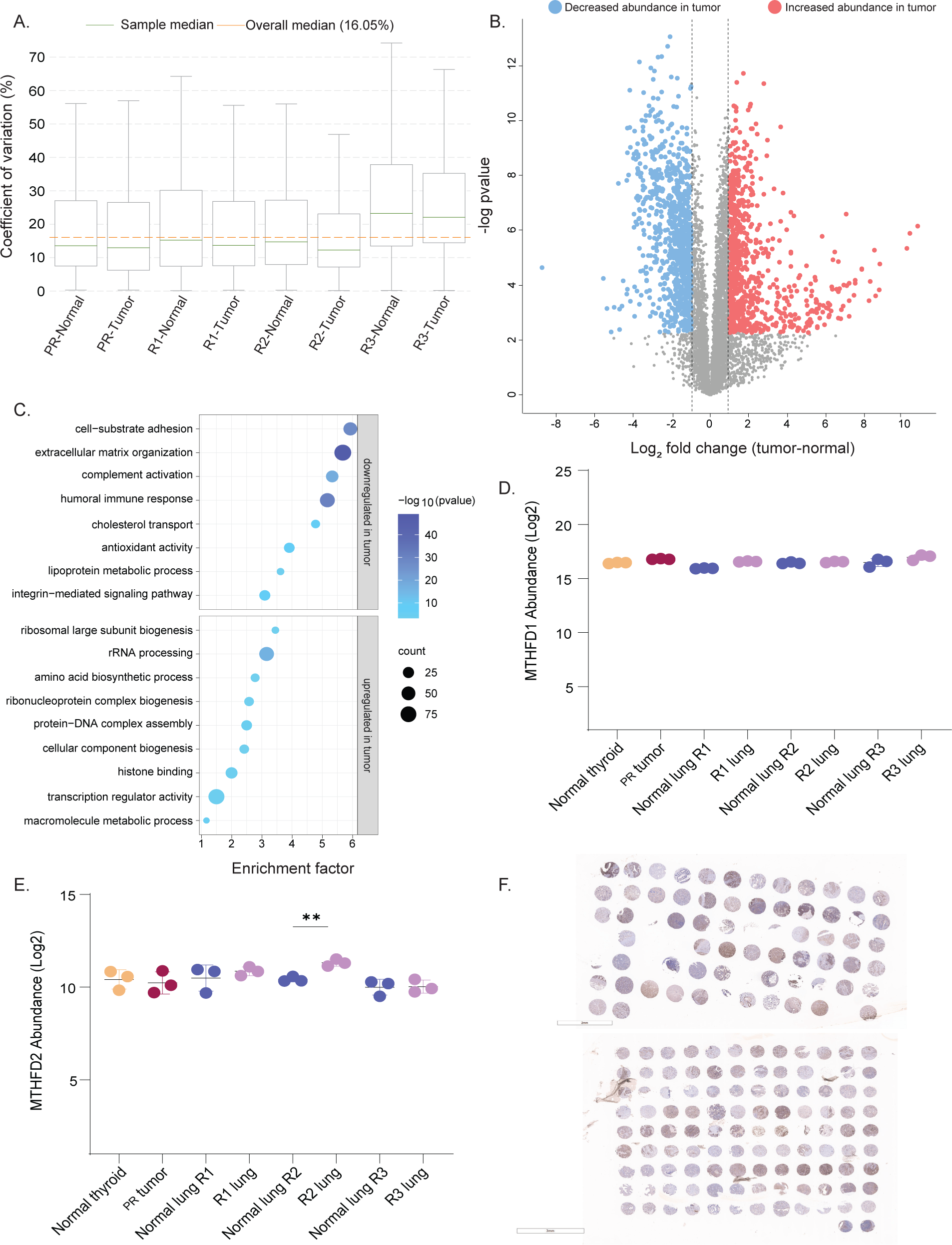
Pathway analysis of the primary tumor and lung metastases as well as corresponding normal tissues at all time points. **A.** Coefficients of variation of protein quantifications across tissue samples collected at different time points. **B.** Volcano plot showing the global analysis of proteome perturbation between the tumor regions and adjacent normal regions. Proteins significant with student’s t-test at 0.01 FDR and with log2 fold change >1 are highlighted in red for upregulation in tumor and in blue for downregulation in tumor. **C.** Pathway enrichment analysis of t-test significant proteins against GO terms using Fisher-exact test, FDR 0.05% show enrichment of proteins involved in transcription regulation, histone binding, protein-DNA complex assembly, amino acid biosynthesis in the primary and lung metastases. Proteins involved in extracellular matrix assembly, cell-substrate adhesion, cholesterol transport, antioxidant activity, and complement activation pathways were enriched in proteins with low abundance in the tumors. **D-E.** Abundance of Methylenetetrahydrofolate dehydrogenases MTHFD1 **(D)** and MTHFD2 **(E)** in the primary tumor and in the lung relapses showing increased abundance of MTHFD2 in the R2. **F.** SHMT2 IHC in TMA 15-012 (top) and TMA 21-003 (bottom). **-pvalue <0.01. Coefficients of variation of protein quantifications across different time points.

**Suppl Figure 2:**
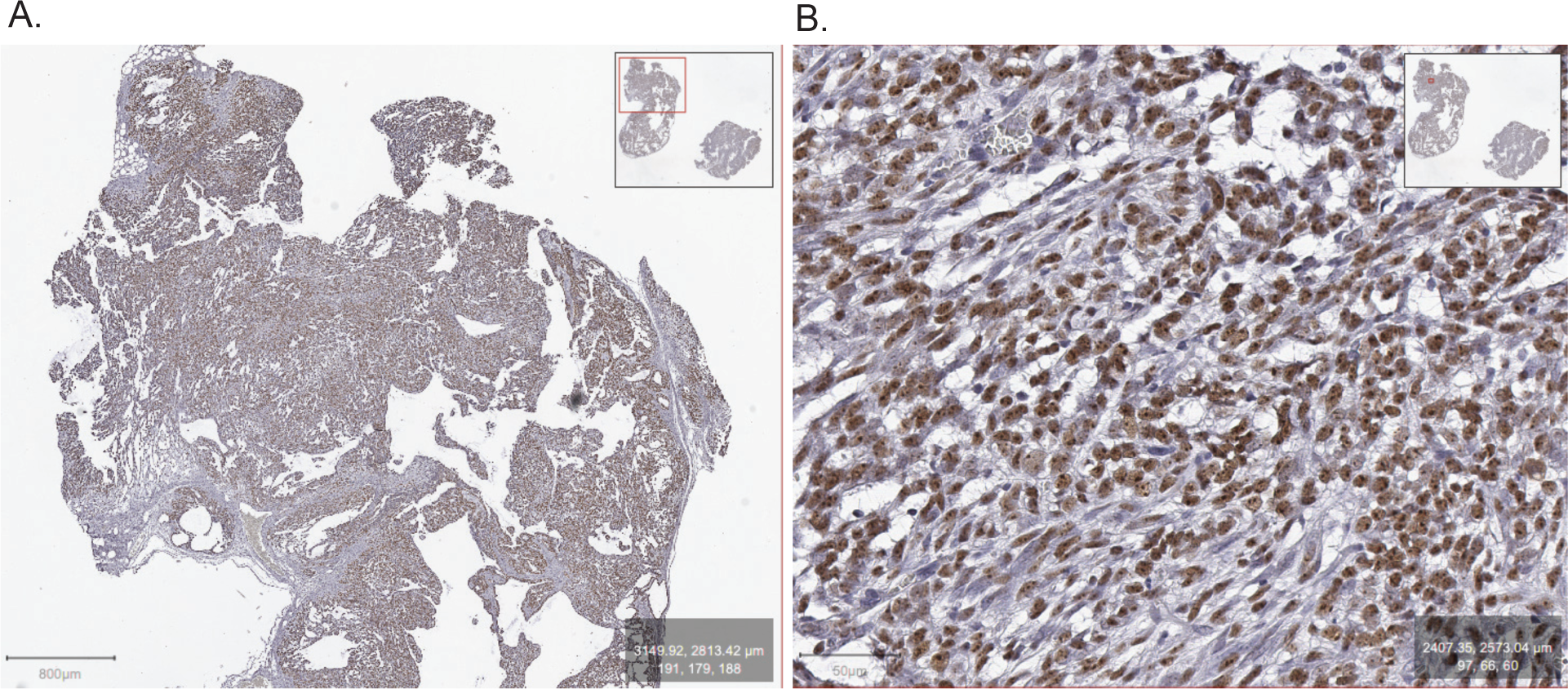
Characterization of SETTLE patient derived mouse xenograft (NSG-PDX) model. Shown are digitally scanned images of FFPE tissue sections isolated from a SETTLE NSG-PDX (*in vivo* passage 1) and stained with a human-specific nucleolin antibody (brown) confirming the presence of human tumor cells. Tissues were counterstained with Toludine blue (blue). Scale bar, 800 µm (left); 50 µm (right).

**Suppl Figure 3:**
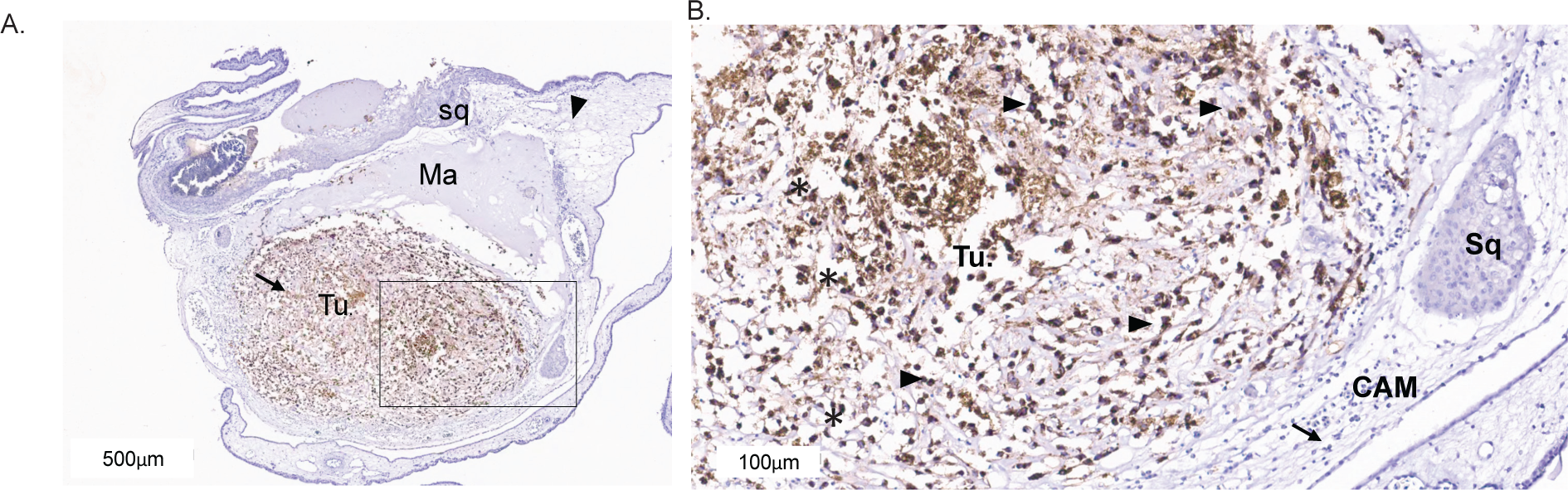
Characterization of SETTLE patient derived chorioallantoic membrane xenograft (CAM-PDX) model. **A.** Low-power magnification of the CAM showing a cellular mass (Arrow) surrounded by the Matrigel (Ma). Squamous metaplasia (Sq) of the chorion is noted on the top of the CAM (arrowhead) and the tumor is encapsulated in the center (Tu.). The tumor in the center is well demarcated as indicated by IHC for a human-specific marker, LAMP1. **B.** Magnified image of CAM-PDX consists of largely discohesive cells with mild to moderate pleomorphism and hyperchromasia with some cells that are oval-to-spindled in appearance with irregular nuclear membranes and scant cytoplasm. Mitoses (Mi) are regularly identified. LAMP1 IHC shows strong cytoplasmic staining in tumor (tu. & arrowheads) cells but not in the CAM membrane (black arrow) or in the squamous metaplasia (sq).

**Suppl Figure 4:**
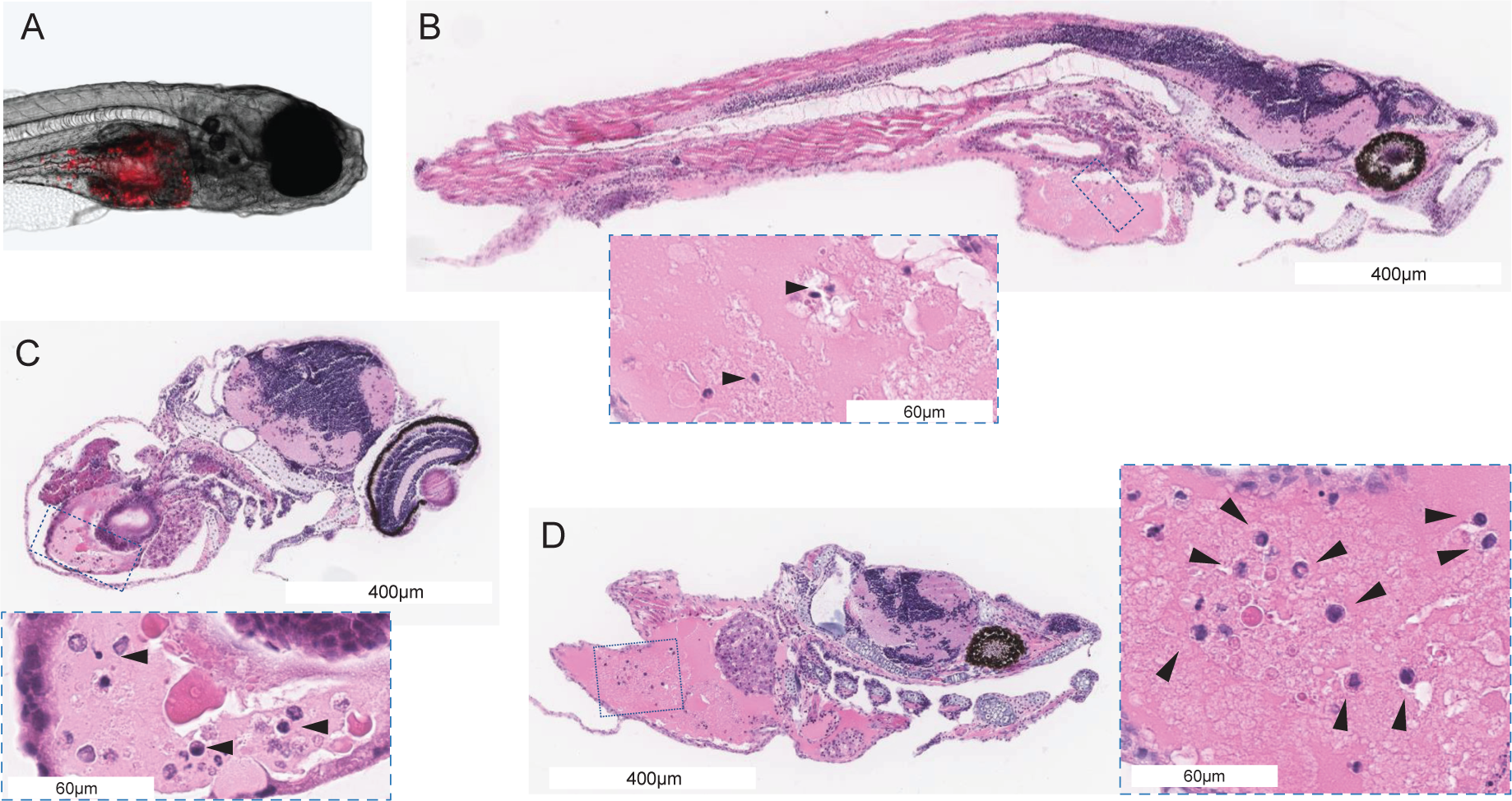
Characterization of zebrafish-PDX model of SETTLE. **A.** Larval zebrafish xenografted with fluorescently labeled NSG-PDX-derived SETTLE tumor cells (red) at 4 days post-implantation (dpi) into the yolk sac. **B, C, D.** Representative H&E-stained FFPE sections of zebrafish-PDX showing SETTLE tumor cells within the yolk sac (arrowheads).

**Suppl Figure 5:**
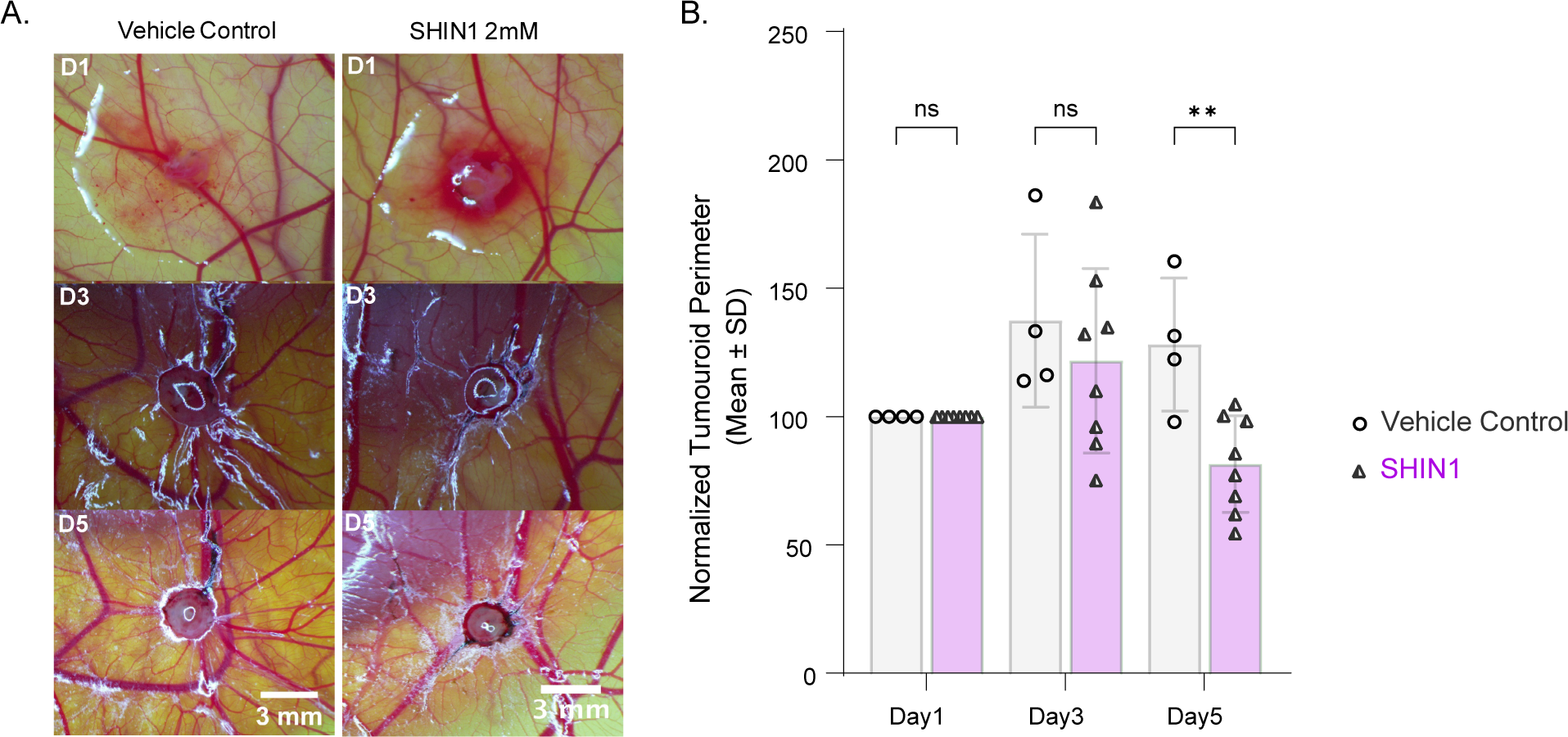
Pre-clinical targeting the one-carbon metabolic pathway in SETTLE tumor using PDX model. **A.** Representative images of SETTLE CAM-PDX over five days with and without (Vehicle Control) treatment with SHIN1. **B.** Measurements of CAM-PDX tumoroid perimeters normalized to Day 1 for untreated and treatment with SHIN1 (**P<0.01).

**Suppl Figure 6:**
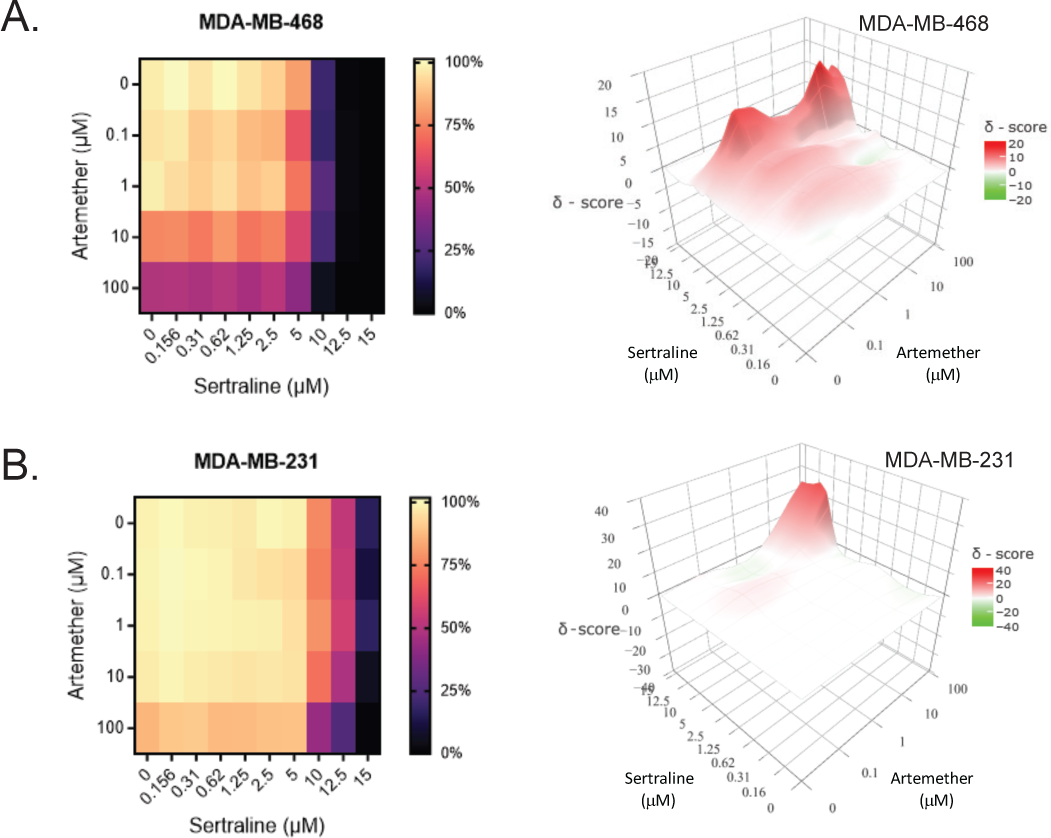
*In vitro* viability assays of breast cancer cell lines (A) MDA-MB-468 and (B) MDA-MB 231 subjected to treatment with sertraline and/or artemether shown as heat map viability plots (left) and the Highest Single Agent combinatorial drug synergy plots (d-score >10 indicative of synergistic effects) (right).

## References

1. K. P. S. Langenberg, E. J. Looze, J. J. Molenaar, The Landscape of Pediatric Precision Oncology: Program Design, Actionable Alterations, and Clinical Trial Development. Cancers (Basel) 13, (2021).

2. K. T. Vo, D. W. Parsons, N. L. Seibel, Precision Medicine in Pediatric Oncology. Surg Oncol Clin N Am 29, 63–72 (2020).

3. K. Strzebonska et al., Risk and Benefit for Targeted Therapy Agents in Pediatric Phase II Trials in Oncology: A Systematic Review with a Meta-Analysis. Target Oncol 16, 415–424 (2021).

4. M. Waligora et al., Risk and surrogate benefit for pediatric Phase I trials in oncology: A systematic review with meta-analysis. PLoS Med 15, e1002505 (2018).

5. R. J. Deyell et al., Whole genome and transcriptome integrated analyses guide clinical care of pediatric poor prognosis cancers. Nat Commun 15, 4165 (2024).

6. S. S. Nishizuka, G. B. Mills, New era of integrated cancer biomarker discovery using reverse-phase protein arrays. Drug Metab Pharmacokinet 31, 35–45 (2016).

7. L. W. Wahjudi et al., Integrating proteomics into precision oncology. Int J Cancer 148, 1438–1451 (2021).

8. Canadian Cancer Statistics 2023 (0835–2976, 2023).

9. J. K. McLoone et al., A Scoping Review Exploring Access to Survivorship Care for Childhood, Adolescent, and Young Adult Cancer Survivors: How Can We Optimize Care Pathways? Adolesc Health Med Ther 14, 153–174 (2023).

10. J. R. Downing et al., The Pediatric Cancer Genome Project. Nat Genet 44, 619–622 (2012).

11. C. McLeod et al., St. Jude Cloud: A Pediatric Cancer Genomic Data-Sharing Ecosystem. Cancer Discov 11, 1082–1099 (2021).

12. P. Berlanga et al., The European MAPPYACTS Trial: Precision Medicine Program in Pediatric and Adolescent Patients with Recurrent Malignancies. Cancer Discov 12, 1266–1281 (2022).

13. C. M. van Tilburg et al., The Pediatric Precision Oncology INFORM Registry: Clinical Outcome and Benefit for Patients with Very High-Evidence Targets. Cancer Discov 11, 2764–2779 (2021).

14. A. C. Uzozie et al., PDX models reflect the proteome landscape of pediatric acute lymphoblastic leukemia but divert in select pathways. J Exp Clin Cancer Res 40, 96 (2021).

15. M. Hidalgo et al., Patient-derived xenograft models: an emerging platform for translational cancer research. Cancer Discov 4, 998–1013 (2014).

16. D. Ribatti, The chick embryo chorioallantoic membrane (CAM). A multifaceted experimental model. Mech Dev 141, 70–77 (2016).

17. P. Kunz, A. Schenker, H. Sahr, B. Lehner, J. Fellenberg, Optimization of the chicken chorioallantoic membrane assay as reliable in vivo model for the analysis of osteosarcoma. PLoS One 14, e0215312 (2019).

18. P. Pawlikowska et al., Exploitation of the chick embryo chorioallantoic membrane (CAM) as a platform for anti-metastatic drug testing. Sci Rep 10, 16876 (2020).

19. A. C. Tufan, N. L. Satiroglu-Tufan, The chick embryo chorioallantoic membrane as a model system for the study of tumor angiogenesis, invasion and development of anti-angiogenic agents. Curr Cancer Drug Targets 5, 249–266 (2005).

20. E. T. Keller, J. M. Murtha, The use of mature zebrafish (Danio rerio) as a model for human aging and disease. Comp Biochem Physiol C Toxicol Pharmacol 138, 335–341 (2004).

21. Q. Tang et al., Optimized cell transplantation using adult rag2 mutant zebrafish. Nat Methods 11, 821–824 (2014).

22. G. Recondo, Jr., N. Busaidy, J. Erasmus, M. D. Williams, F. M. Johnson, Spindle epithelial tumor with thymus-like differentiation: A case report and comprehensive review of the literature and treatment options. Head Neck 37, 746–754 (2015).

23. J. R. Grushka et al., Spindle epithelial tumor with thymus-like elements of the thyroid: a multi-institutional case series and review of the literature. J Pediatr Surg 44, 944–948 (2009).

24. T. M. Stevens et al., Spindle Epithelial Tumor with Thymus-Like Differentiation (SETTLE): A Next-Generation Sequencing Study. Head Neck Pathol 13, 162–168 (2019).

25. C. L. Matheson, G. K. Blair, J. Bush, Spindle Epithelial Tumor with Thymus-Like Differentiation (SETTLE): A Case Report. Fetal Pediatr Pathol 38, 399–405 (2019).

26. G. D. Barnabas, V. Goebeler, J. Tsui, J. W. Bush, P. F. Lange, ASAP horizontal line Automated Sonication-Free Acid-Assisted Proteomes horizontal line from Cells and FFPE Tissues. Anal Chem 95, 3291–3299 (2023).

27. F. Luongo et al., PTEN Tumor-Suppressor: The Dam of Stemness in Cancer. Cancers (Basel) 11, (2019).

28. A. C. Lorentzian et al., Targetable lesions and proteomes predict therapy sensitivity through disease evolution in pediatric acute lymphoblastic leukemia. Nat Commun 14, 7161 (2023).

29. P. S. Ward, C. B. Thompson, Metabolic reprogramming: a cancer hallmark even warburg did not anticipate. Cancer Cell 21, 297–308 (2012).

30. Y. C. Chan et al., Overexpression of PSAT1 promotes metastasis of lung adenocarcinoma by suppressing the IRF1-IFNgamma axis. Oncogene 39, 2509–2522 (2020).

31. T. De Marchi et al., Phosphoserine aminotransferase 1 is associated to poor outcome on tamoxifen therapy in recurrent breast cancer. Sci Rep 7, 2099 (2017).

32. M. Feng et al., An integrated pan-cancer analysis of PSAT1: A potential biomarker for survival and immunotherapy. Front Genet 13, 975381 (2022).

33. M. Y. Huang et al., Phosphoserine phosphatase as a prognostic biomarker in patients with gastric cancer and its potential association with immune cells. BMC Gastroenterol 22, 1 (2022).

34. L. Liao et al., Upregulation of phosphoserine phosphatase contributes to tumor progression and predicts poor prognosis in non-small cell lung cancer patients. Thorac Cancer 10, 1203–1212 (2019).

35. B. Liu et al., Overexpression of Phosphoserine Aminotransferase 1 (PSAT1) Predicts Poor Prognosis and Associates with Tumor Progression in Human Esophageal Squamous Cell Carcinoma. Cell Physiol Biochem 39, 395–406 (2016).

36. S. Bernhardt et al., Proteomic profiling of breast cancer metabolism identifies SHMT2 and ASCT2 as prognostic factors. Breast Cancer Res 19, 112 (2017).

37. S. L. Geeraerts et al., Repurposing the Antidepressant Sertraline as SHMT Inhibitor to Suppress Serine/Glycine Synthesis-Addicted Breast Tumor Growth. Mol Cancer Ther 20, 50–63 (2021).

38. L. Ji, Y. Tang, X. Pang, Y. Zhang, Increased Expression of Serine Hydroxymethyltransferase 2 (SHMT2) is a Negative Prognostic Marker in Patients with Hepatocellular Carcinoma and is Associated with Proliferation of HepG2 Cells. Med Sci Monit 25, 5823–5832 (2019).

39. X. Jiang, et al., Repurposing sertraline sensitizes non-small cell lung cancer cells to erlotinib by inducing autophagy. JCI Insight 3, (2018).

40. A. Irmisch et al., The Tumor Profiler Study: integrated, multi-omic, functional tumor profiling for clinical decision support. Cancer Cell 39, 288–293 (2021).

41. J. J. Tentler et al., Patient-derived tumour xenografts as models for oncology drug development. Nat Rev Clin Oncol 9, 338–350 (2012).

42. C. Rebelo de Almeida et al., Zebrafish xenografts as a fast screening platform for bevacizumab cancer therapy. Commun Biol 3, 299 (2020).

43. J. Q. Wu et al., Patient-derived xenograft in zebrafish embryos: a new platform for translational research in gastric cancer. J Exp Clin Cancer Res 36, 160 (2017).

44. F. Petralia et al., Integrated Proteogenomic Characterization across Major Histological Types of Pediatric Brain Cancer. Cell 183, 1962–1985 e1931 (2020).

45. P. F. Lange, O. Schilling, P. F. Huesgen, Positional proteomics: is the technology ready to study clinical cohorts? Expert Rev Proteomics 20, 309–318 (2023).

46. D. Akhoundova, M. A. Rubin, Clinical application of advanced multi-omics tumor profiling: Shaping precision oncology of the future. Cancer Cell 40, 920–938 (2022).

